# A Tunable Flat-Jet Hydro-Debridement Device: Clinical Feasibility for Soft Tissue Wound Management

**DOI:** 10.64898/2026.09.01.26361120

**Authors:** Arkadeep Datta, Rudrajit Majumder, Indranil Biswas, Ranjan Ganguly, Apurba Kumar Santra, Sourav Sarkar, Manas Kumar Gumta, Subhasish Sarkar

**Affiliations:** Advanced Materials Research and Application Laboratory, Department of Power Engineering, Jadavpur University (Salt Lake campus), West Bengal – 700106; Department of Mechanical Engineering, Jadavpur University, West Bengal – 700032; Department of General Surgery, Bankura Sammilani Medical College & Hospital. Lokepur, PO- Kenduadihi, Dist- Bankura, West Bengal – 722102; Department of General Surgery, College of Medicine &Sagore Dutta Hospital (CMSDH), Kamarhati, West Bengal - 700058

**Keywords:** Adjustable flow, Affordable, Hydro-debridement system, Pressurized irrigation, Ulcer, Wound healing

## Abstract

**Background:** Chronic wounds, ulcers, and lacerations require staged debridement and irrigation to promote healing. However, conventional techniques of debridement, such as surgical, chemical, or autolytic, struggle to fully remove residual necrotic tissue, slough, and unhealthy granulation from wound sites, especially when lodged within wound clefts and cavities, and in wounds with exposed structures. This promotes polymicrobial biofilms, delays wound closure, and causes significant discomfort with increased morbidity.

**Objective:** To demonstrate the feasibility of using an indigenously developed tunable flat-jet hydro-debridement device (presently termed as CleanseJet), a frugal wound debridement system designed for deployment in resource-constrained clinical settings.

**Methods:** An open-label, interventional, single-centre, parallel-group pilot randomized controlled trial was conducted to clinically evaluate an indigenously developed tunable flat-jet hydro-debridement device in patients with wounds of varied aetiology. The device provided adjustable spray impact force and coverage area tailored to wound characteristics. Outcomes were compared with a control group receiving standard wound care alone, with time to complete granulation serving as the primary healing endpoint. Outcomes were compared with a control cohort receiving standard of care alone.

**Results:** The removal of loose devitalized tissue, slough, and biofilms from the wound bed improved the healing, which were monitored using the SINBAD scoring system. No adverse events were reported, supporting the feasibility and safety of CleanseJet.

**Conclusion:** While commercial hydro-debridement systems are effective, they are often costly, rely on disposable components, and require specialized training. In contrast, CleanseJet provides a low-cost, easy-to-use alternative that can be operated with minimal training, making it suitable for broader clinical use without observed adverse effects.

**Graphical Abstract:** 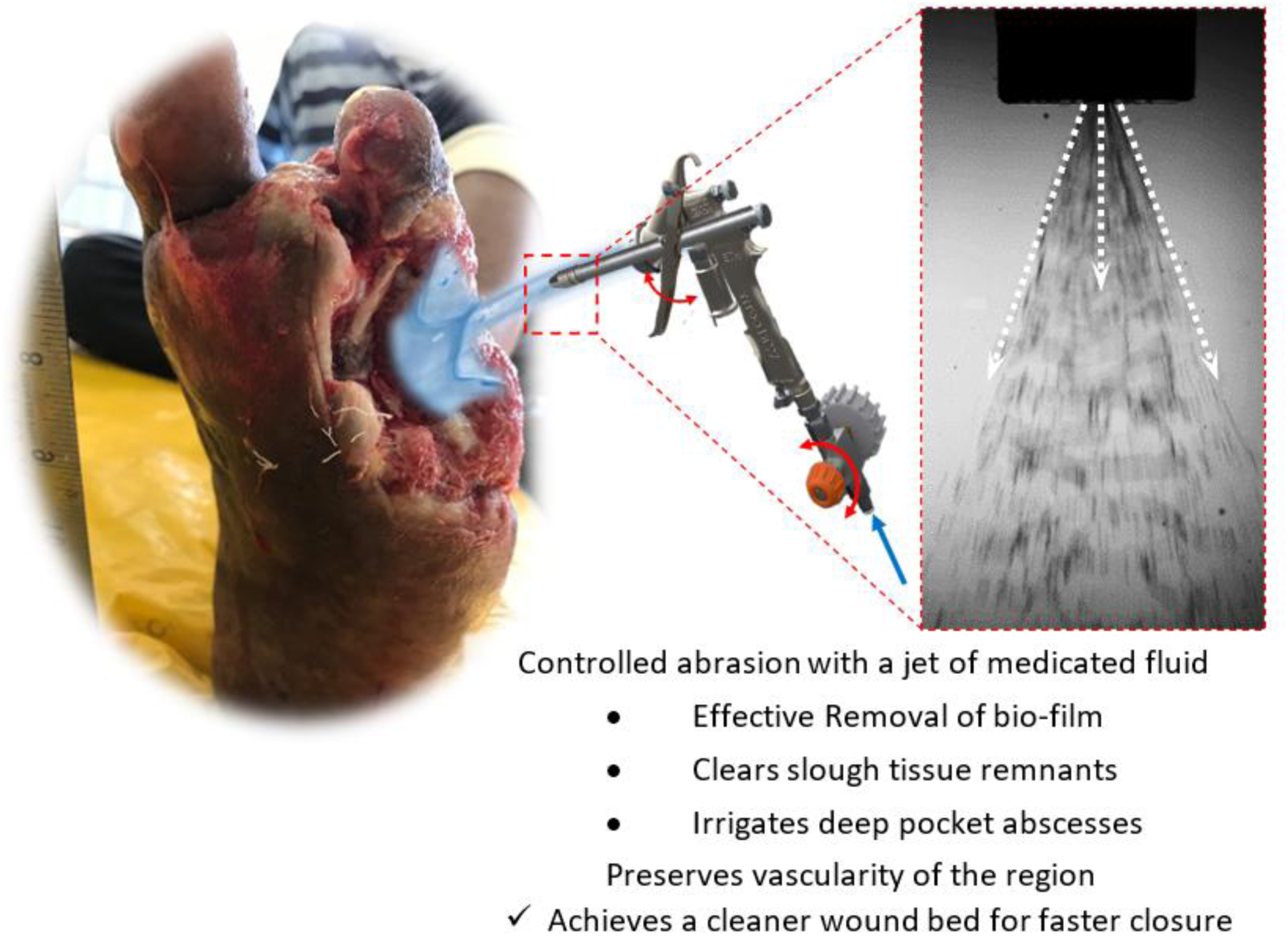

## Introduction

Managing chronic ulcers remains a major clinical challenge, particularly among individuals with diabetes, where the lifetime risk approaches 15%.^1^ These wounds are frequently complicated by neuropathy, ischemia, necrotic tissue accumulation, fibrin deposits, and biofilms, all of which delay healing and increase the risk of spreading cellulitis and amputation.^2^ Effective wound bed preparation, as emphasized by the TIME framework,^3^ requires timely removal of nonviable tissue.^4^ The standard of care (SoC) in such cases often includes surgical debridement, mechanical debridement with wet-to-dry gauze,^5^ which may be painful and nonselective, potentially traumatizing the underlying viable tissues. While chemical^6^ and autolytic^7^ methods are slower and often require repeated clinical visits, leading to increased treatment duration, impacting both cost^8^ and quality of life of the patient.^9^

Non-conventional wound management techniques like negative pressure wound therapy (NPWT)^10^ or other adjunctive techniques are available, but they may not be practical in resource-constrained settings with a high patient-to-care-provider ratio. Alternatively, hydro-debridement provides a targeted and minimally invasive solution – using pressurized saline jets to flush out loose necrotic tissue^11, 12^ with pressurized medicated fluids (0.9% NS), lowering bacterial load^13^ while minimizing tissue trauma^14^ due to mechanical-abrasion, usually practised in the standard of care (SoC).^15^ While commercial systems like Debritom™ (Amtech Medaxis), VERSAJET™ hydroscalpel (Smith & Nephew),^16^ Erbe Elektromedizin GmbH,^17^ hydrocision^18^ and others have shown effectiveness in debriding chronic ulcers,^19^ their effectiveness in removing high necrotic loads, thin polymicrobial biofilms, and irrigation of deep cavities or complex wound geometries^20^ are limited. Additionally, these systems are often expensive, dependent on disposable components, and may have limited flexibility for use in debriding hard eschars, which still require surgical intervention.

## Clinical implication

CleanseJet, a low-cost hydro-debridement device with tunable saline water spray, is designed to support controlled hydro-debridement and irrigation, particularly in wounds with deep cavities following surgical clearance of dense necrosis. The scope of the present pilot study is limited to wounds containing loose necrotic tissue and slough and did not include evaluation of hard or firmly adherent necrotic tissue. Within this scope, the device demonstrated effective selective debridement while preserving viable tissue, particularly in deep cavity wounds, without any noticeable device-related adverse effects.

At the bedside, this approach may provide clinicians with a more controlled and potentially less traumatic alternative to conventional mechanical debridement for routine wound bed preparation. Its simple design and minimal training requirements may allow integration into busy outpatient clinics and resource-limited settings where access to advanced and expensive hydro-debridement platforms are restricted. Although larger comparative studies are warranted, early feasibility findings suggest that CleanseJet may help streamline wound cleansing while maintaining tissue integrity.

## Materials and methods

### Study design, subjects, and interventions

A 20-week open-label pilot study was conducted at a tertiary referral public hospital cum medical college to evaluate the clinical feasibility and safety of the hydro-debridement device. 50 patients (all aged over 18 years) presenting with a range of ulcer types were screened for eligibility and subsequently randomized into either the intervention group (hydro-debridement device plus standard wound care) or the control group (standard wound care alone). Moribund patients and those with ulcers complicated by lymphedema were excluded. The number of days taken to achieve complete healthy granulation tissue were considered as end time point. Outcomes were compared with a control cohort receiving standard of care only.

### Ethical Considerations

The study was approved by the Institutional Ethics Committee and conducted in accordance with institutional guidelines. It forms part of a larger interventional clinical trial registered with Clinical Trials Registry – India (CTRI/2025/08/092962). Reporting follows the *CONSORT Extension for Pilot and Feasibility Trials (2016)*. Written informed consents were obtained from all participants, and confidentiality was maintained throughout.

### Wound Assessment and Monitoring

Given the heterogeneity of wound types and patient comorbidities, standardized classification systems were used to support structured documentation. WAGNER grading^21^ was applied at baseline, and the SINBAD classification^22^ was used for serial follow-up assessments due to its greater specificity^23^ (see **Section S1.0** in electronic supplementary information (*ESI*) for details). Weekly evaluations documenting wound characteristics, healing progression, and any adverse events, including excessive bleeding or unintended tissue injury was assessed to determine procedural safety and feasibility (see section below and **Section S4.0** in the *ESI* for details). The wounds were evaluated clinically, and all interventional participants, after each hydro-debridement session with CleanseJet, received SoC while the control group received only SoC.^24^ The SoC protocol, applied to both groups, include wound bed preparation, topical antimicrobial therapy, appropriate dressings, and multicomponent compression therapy wherever indicated. Participants were evaluated weekly in the outpatient setting of the public hospital.

### The Wagner grading

The Wagner grading, as adopted in the study and elucidated in **Figure S1** in the ESI document, clinically assesses ulcer-depth and involvement of deeper structures using the following grades:

- **Wagner Grade 0:** Skin is intact with no open lesion or a pre-ulcerative lesion – may have a deformity or cellulitis
- **Wagner Grade 1:** Partial- or full-thickness ulcer (superficial ulcer)
- **Wagner Grade 2:** Deep ulcer extended to ligament, tendon, joint capsule, bone, or deep fascia without abscess or osteomyelitis (OM)
- **Wagner Grade 3:** Deep abscess, OM, or joint sepsis
- **Wagner Grade 4:** Partial-foot gangrene
- **Wagner Grade 5:** Whole-foot gangrene

Assessing the wound conditions using the WAGNER grade, at the patient’s initial visit facilitated a quicker evaluation and recruitment into the study. Additionally, this grading allowed for a prompt determination of the required necrotic tissue resection and the implementation of appropriate medical procedures during the first consultation.

### The SINBAD classification system

The SINBAD classification is additionally adopted for its greater specificity and detail. The SINBAD system evaluates six clinical parameters as follows, resulting a score between 0 and 6:

- *Ulcer **S**ite*: Forefoot (0) and midfoot/hindfoot (1)
- ***I****schemia*: Intact blood flow (0) and evidence of ischemia [neither pulse palpable with signs of reduced tissue perfusion, with or without gangrene] (1)
- ***N****europathy*: Absent (0) or present (1), assessed clinically
- ***B****acterial infection*: Absent (0) or present (1), based on clinical signs of soft tissue or bone infection^4^
- ***A****rea*: ≤1 cm² (0) or >1 cm² (1), determined by multiplying the two maximum dimensions at right angles
- ***D****epth*: Superficial (0) or deep, reaching tendon, periosteum, joint capsule, or bone (1)

The individual grades are summed to produce a SINBAD score ranging from 0 to 6.

In the absence of a universally accepted wound grading system applicable to wounds of varied etiologies, wound status was assessed using the Wagner and SINBAD scoring systems across the study cohort, rather than limiting their use to diabetic foot ulcers. Therefore, the use of Wagner and SINBAD scores for non-diabetic foot wounds may not have fully captured wound severity or healing progression in those cases.

### The CleanseJet system and its deployment

The CleanseJet system (see **Figure 1A**) is a portable hydro-debridement platform consisting of a handheld wand-like spray-device, a pressurization system, and a saline sump. The spray-device is fitted with a flat fan nozzle (Spraying Systems Co., Part No. H 1/8 DT–SS-25-01)^25^ at the distal end to deliver a controlled, pressurized sheet of 0.9% normal saline (NS). The nozzle is rated at a 25° fan angle, with a fluid pressure of ∼ 3 bar at the nozzle entry. The pump delivers ∼ 1.9 L/min NS at a maximum operating pressure of ∼ 5 bar. Droplets generated by the spray, upon impacting the wound surface, enable selective removal of loosely adherent devitalized tissue while minimizing the risk of deep tissue penetration.

**Figure 1.**
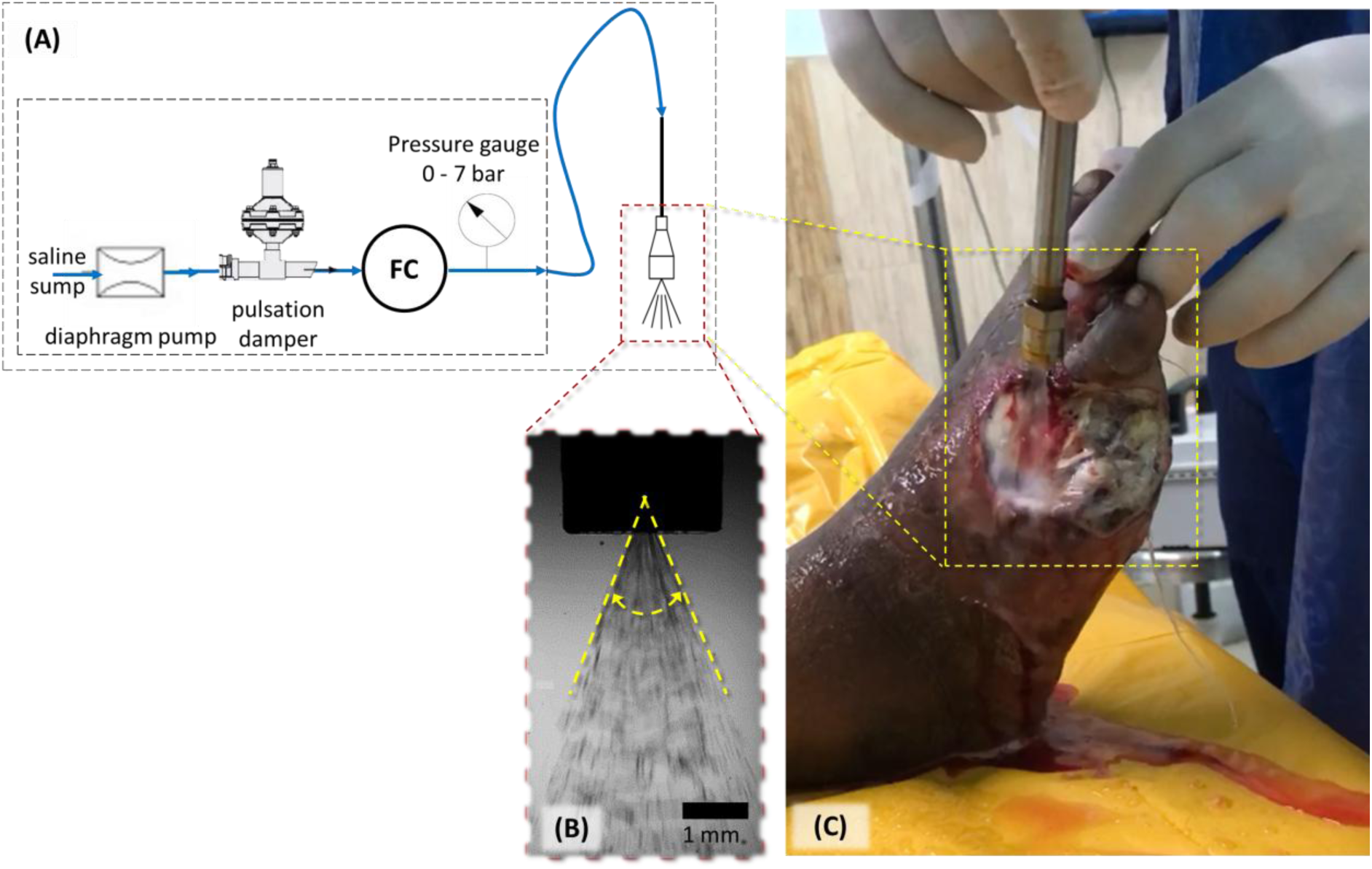
**(A)** Schematic of the hydraulic circuit of the tuneable pressure saline spray system, CleanseJet. The system employs a positive displacement pump to pressurize medical-grade saline (0.9% NS), producing a flat spray with a cone angle of ∼ 45° and a flow rate of up to ∼ 1.2 L/min **(B)** Image illustrating the spray characteristics (cone angle) from the nozzle tip. **(C)** Hydro-debridement on a participant with amputated fourth and fifth digits of the right foot. The image shows a well-irrigated and cleaned wound bed. The dotted white arrows denote the flow of 0.9% NS over the wound surface. Any loose tissue debris and biofilm washed away by the fluid stream drains by gravity (see the green arrow).

In practice, the saline jet functions as a noncontact mechanical curette (pseudo-scalpel), facilitating controlled wound bed cleansing, while assuring that deep penetration into the underlying tissue is avoided.

Made from stainless steel grades 316 and 303, respectively, the device is autoclavable, permitting repeated sterilization and reuse. This inherently enhances cost-effectiveness and reduces medical waste. The slim, wand-like configuration supports precise positioning, enabling clinicians to access wound clefts, undermined areas, and irregular surfaces with improved control. The console, connected to the wand via a high-pressure hose, houses a flow control valve (needle valve) (FC in **Figure 1A**), a positive displacement pump (Model No. RQS300G), and a pressure gauge.

**Figure 1A** – **Inset** shows deployment of the CleanseJet system by paramedics for debridement on a patient with diabetic foot ulcer (DFU) in the out-patient department of a hospital. For the current study, the flow rate is typically maintained at approximately 1.2 L/min (spray fan angle ∼ 45°, measured optically; **Figure 1B**), with an operating pressures substantially lower than those used in commercial hydro-surgery platforms.^20^ Spray impact can be further adjusted by altering the stand-off distance between the nozzle and the wound surface,^26^ providing an additional layer of procedural control. A pulsation damper, installed in series between the pump and flow control valve, dampens any pulsations in the spray – ensuring consistent spray delivery during use.

In clinical application (**Figure 1C**), pressurized saline is directed over the wound surface following surgical debridement when indicated. The system enables simultaneous hydro-debridement, cavity lavage, and wound bed irrigation within a single workflow. Effluent and debris are removed by gravity drainage. Supplementary videos *ESI* **Movies 1** and **2** demonstrate its use in a trauma and a burn wound respectively, where adherent slough was effectively cleared, and eschar softened to facilitate gentle removal. Post-irrigation findings typically include a visibly cleaner wound bed with mild reactive erythema consistent with early inflammatory healing

The functionality of the CleanseJet system is compared to the SoC and further through an account of its application on wounds with different wound aetiology, site, and size below.

### Operation of the CleanseJet system

**Figure 2** and the supplementary materials compare the performance of CleanseJet with conventional irrigation techniques. Standard syringe irrigation or free-flow saline application often provides limited penetration into wound recesses and may require higher fluid volumes. In contrast, CleanseJet delivers focused, directional irrigation with reduced splash and improved visualization of the wound bed.

**Figure 2:**
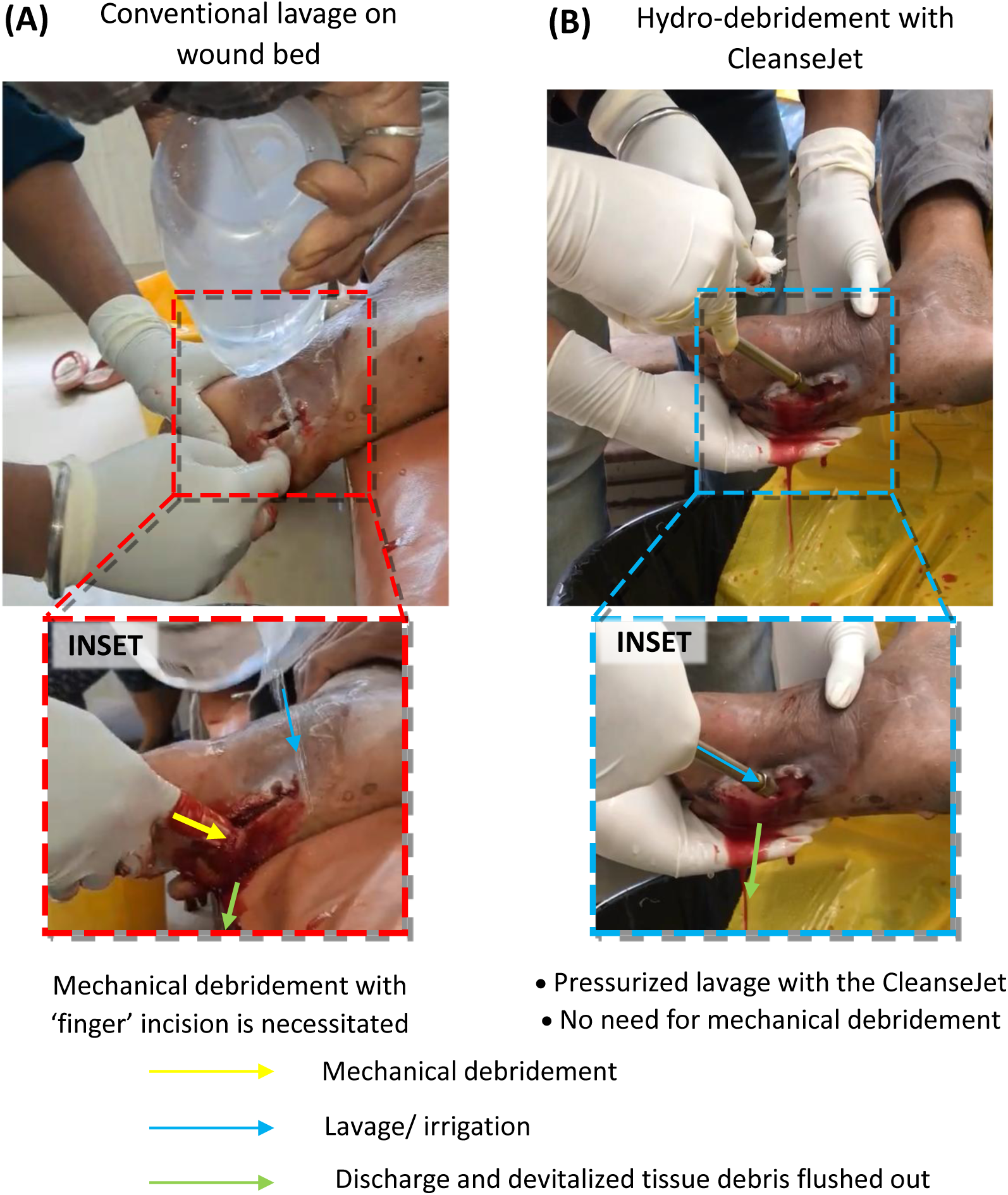
The participant presents with acute diabetes and a swollen feet with a bulge in footpad. Clinical observations indicated the presence of loculations of discharge present in a cavity in the planter region. An incision is made along the axis of the sole as seen in **(A)** with an intension to lavage the cavity off any discharge in the region. INSET: Such conventional methods of irrigation at times fails to evacuate all discharge and mechanical debridement with ‘finger’ incision in the area is necessitated. Such methods risks damaging tendons, arteries and other structures present in the area. **(B)** Pressurized spray of saline with the CleanseJet reaches inside cavities in such difficult-to-reach areas of the wound bed, effectively flushing out devitalized tissue debris and discharges. INSET: Fluid pressure and flow rate are modulated to allow for optimized lavage in the wound-bed. Refer to *ESI* **Movie 3** for further details.

For a participant with a DFU and suspected plantar abscess (**Figure 2A**), surgical exposure revealed loculated purulent collections. Traditional management requires manual disruption of loculations, which may increase the risk to adjacent tendons and neurovascular structures (*ESI* **Movie 3**, left panel). Using CleanseJet (**Figure 2B**; *ESI* **Movie 3**, right panel), the pressurized saline stream dislodged purulent material without direct mechanical manipulation. Spray parameters (flow rate, operating pressure, *etc*.) were adjusted to selectively disrupt nonviable tissues while preserving viable structures. Additional before-after comparisons are provided in **Figure S2** (*ESI*). Across wound types—including DFUs, trauma wounds, and burns—the device facilitates removal of slough, evacuation of purulent pockets, and irrigation of anatomically constrained spaces. No procedure-related adverse events or patient-reported severe discomfort were observed.

### Measures to Minimize Environmental and Cross-Contamination

Hydro-debridement is performed in a designated dressing room under routine institutional infection-control practices. Only one patient is treated at a time, and the treatment area is cleaned and disinfected before subsequent use. Standard wound-care precautions are observed by the treating personnel throughout the procedure. These measures are adopted to minimize the potential risk of splash-or aerosol-mediated dissemination of wound-derived microorganisms. Environmental microbiological monitoring is not undertaken as part of this pilot study.

## Results

### Exploratory clinical outcomes

In a tertiary care hospital, wound care is usually provided till the wound exhibits healthy granulation throughout the wound bed, followed by self-care advice at home until the wound-closure. In this study, we considered complete healthy granulation on the wound bed as the healing event. The clinical outcomes were exploratory in nature and are represented by days taken to achieve complete healthy granulation on the wound bed and wound progression in terms of the SINBAD score. The CONSORT Extension for Pilot and Feasibility Trials (2016) flow diagram is depicted in **Figure 3** in terms of the clinical outcomes, while a qualitative outcome is described in *ESI* **S3.0**.

**Figure 3:**
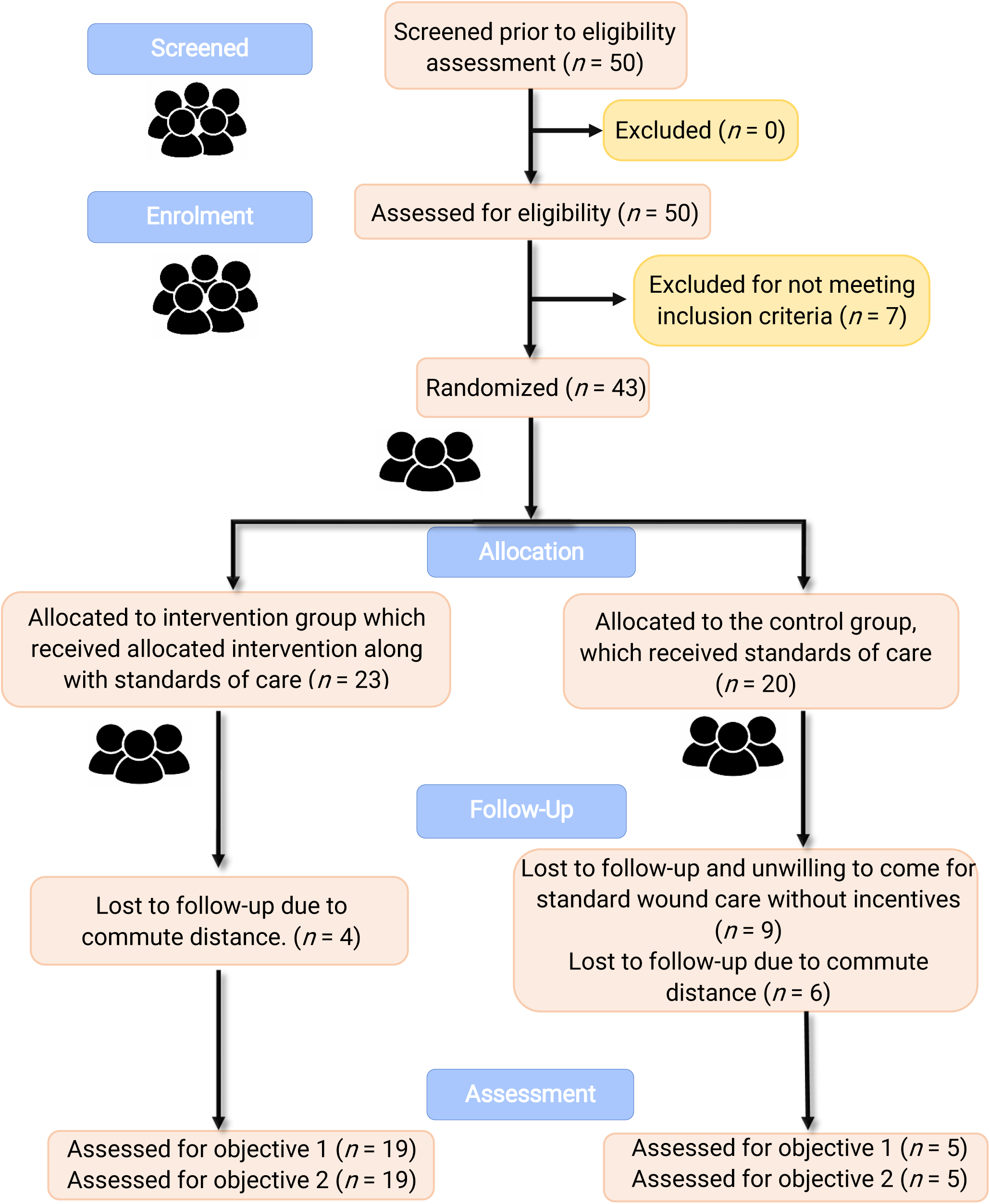
CONSORT Extension for Pilot and Feasibility Trials (2016) flow diagram

### Baseline characteristics

The baseline characteristics of the participants, including age, sex, and Wagner grades of their wounds, are summarised in **Table 1**. Both the intervention and the control group had a mean age of ∼55 years and average Wagner wound grade of ∼2.

**Table 1:** Baseline characteristics: Age, sex, and Wagner grade of wound.

| Characteristic | Intervention (n = 19) | Control (n = 5) | Total (N = 24) |
| --- | --- | --- | --- |
| Age, mean (SD), years | 54.6 (17.4) | 57.2 (14.19) | - |
| Male sex, n (%) | 84 | 60 | - |
| Female sex, n (%) | 16 | 40 | - |
| Wagner grade of wound, mean (SD) | ~2 (~1) | ~2 (~1) | ~2 (~1) |

### Feasibility and outcome measures

50 participants were screened, of whom 43 were randomized. Of these, 24 completed follow-up (intervention: *n* = 19; control: *n* = 5), with complete adherence to the intervention and full data capture for all outcome measures. The intervention group included 19 participants with 20 distinct wound sites treated with CleanseJet hydro-debridement, while the control group received standard of care (SoC) alone. Recruitment, retention, adherence, adverse effects and outcomes are summarised in **Table 2** and **Table 3**, respectively.

**Table 2:**
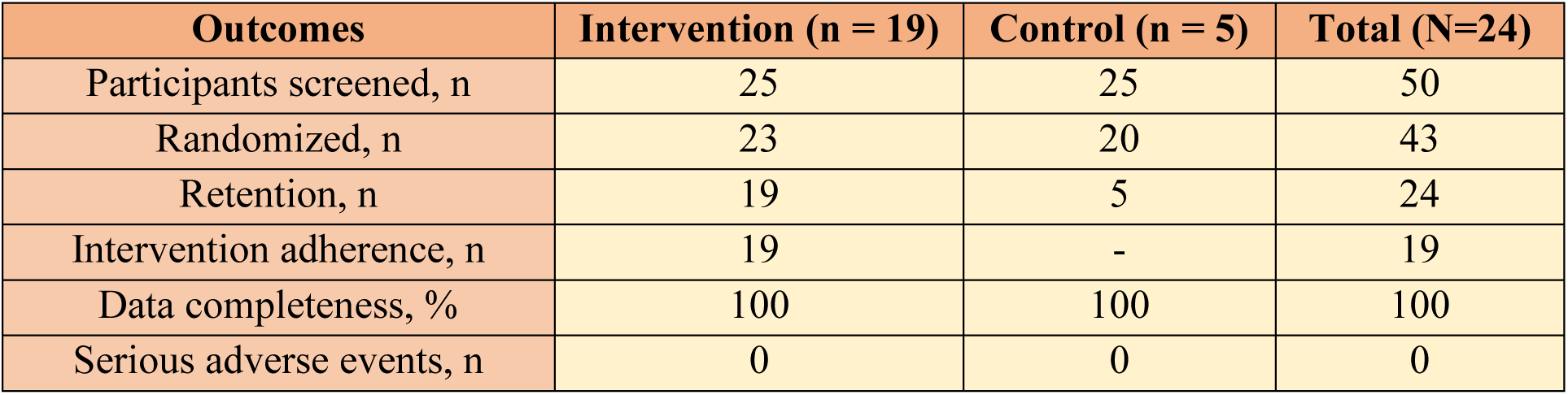
Recruitment, retention, adherence, data completeness, and adverse events.

| Outcomes | Intervention (n = 19) | Control (n = 5) | Total (N=24) |
| --- | --- | --- | --- |
| Participants screened, n | 25 | 25 | 50 |
| Randomized, n | 23 | 20 | 43 |
| Retention, n | 19 | 5 | 24 |
| Intervention adherence, n | 19 | - | 19 |
| Data completeness, % | 100 | 100 | 100 |
| Serious adverse events, n | 0 | 0 | 0 |

**Table 3.** The mean ± SD and CIs of days taken to exhibit complete healthy granulation and final SINBAD score of wounds.

| Outcome | Intervention (n = 19) | Control (n = 5) | Mean difference (95% CI) |
| --- | --- | --- | --- |
| Days taken to exhibit complete healthy granulation, mean (SD) | 48.61 (34.09) | 40.6 (25.91) | 8.01 (-23.15 to 39.17) |
| Final SINBAD score, mean (SD) | 1.94 (1.12) | 1.6 (1.52) | 0.34 (-1.54 to 2.22) |

### Chronological healing progress following the CleanseJet application in three cases of chronic wounds

From the pool of cases given in *ESI* **Section S4.0**, we focused on two cases of chronic ulcers, each differing in wound site, size and aetiology, to demonstrate the effectiveness of CleanseJet in irrigation, lavage and hydro-debridement. ‘Before’–and–‘after’ comparisons in each hydro-debridement session were documented, with weekly monitoring of healing progress using the SINBAD scoring system (see **Figures 4 – 6** and **Figure S4** in *ESI* **Section S4.0**), allowing for a comparative analysis of the wound-bed. Clinical observations, along with SINBAD scores demonstrate a notable reduction in slough and necrotic tissue load within the first few uses of the CleanseJet. The cases are presented in increasing order of wound severity as per WAGNER grading. Participant details and healing durations are summarized in **Table S1** in the *ESI*.

**Figure 4.**
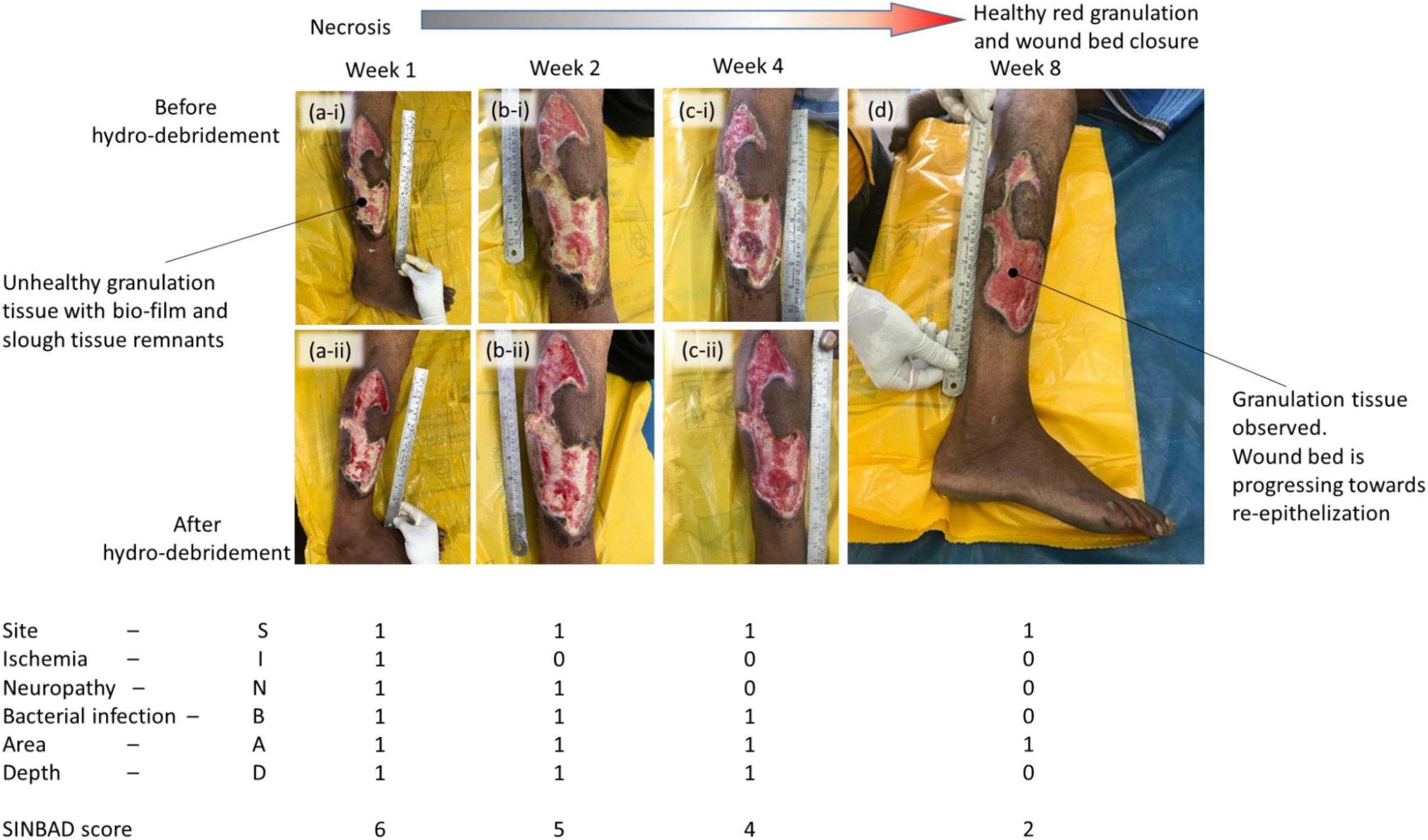
Example of a male participant in his mid-sixties − with type 2 diabetes and severe cellulitis with necrosis on the right lower limb − had undergone surgical debridement and fasciotomy: post-operative care included weekly hydro-debridement with CleanseJet, antibiotics, and diabetic management. Within six weeks, granulation tissue developed across the wound bed, reducing the SINBAD score from 4 to 1, making it conducive for a further recommendation for grafting.

### Case 1: Management of rapidly spreading cellulitis with necrosis

A male participant in the age bracket of 60 – 65 with type 2 diabetes mellitus (T2DM) presented with rapidly spreading cellulitis and necrosis over the lateral aspect of the right lower limb, below the knee. Upon evaluation, surgical debridement with fasciotomy was performed under regional anaesthesia to remove the necrotic tissues and control infection. Post-surgery, bedside dressing revealed extensive slough and purulent discharge in the wound bed, indicative of persistent infection. Wound management included broad-spectrum antibiotics, strict glycaemic control, and weekly CleanseJet hydro-debridement with standard dressings, and was assigned a WAGNER grade of 1. Weekly irrigation with CleanseJet followed with standard medical dressing was applied to promote healing. Each session progressively cleared devitalized tissue, improving SINBAD scores from 4/6 to 1/6 (**Figure 4**). By eight weeks, full granulation was achieved, although complete epithelialization remained incomplete (**Figure 4** d), yielding a SINDBAD score of 1 instead of ’0’). The participant was subsequently referred for reconstructive surgery.

### Case 2: Diabetic foot with wet gangrene and osteomyelitis

A male patient in his early forties with poorly controlled diabetes, and with wet gangrene of the fourth and fifth toes was presented in the out-patient department. Clinical examination and X-ray imaging indicated osteomyelitis of the fifth toe, while X-ray imaging confirmed spread of infection to the fourth toe and with an associated pocket of abscess, consistent with partial foot gangrene (WAGNER grade 4). Immediate surgical intervention, including toe amputation, was performed under regional anaesthesia. Post-surgery, irrigation of the wound bed was performed using the CleanseJet (**Figure 5** a-ii). Care protocols and wound progress documentation consistent with those implemented in Cases 1 and 2 were applied.

**Figure 5:**
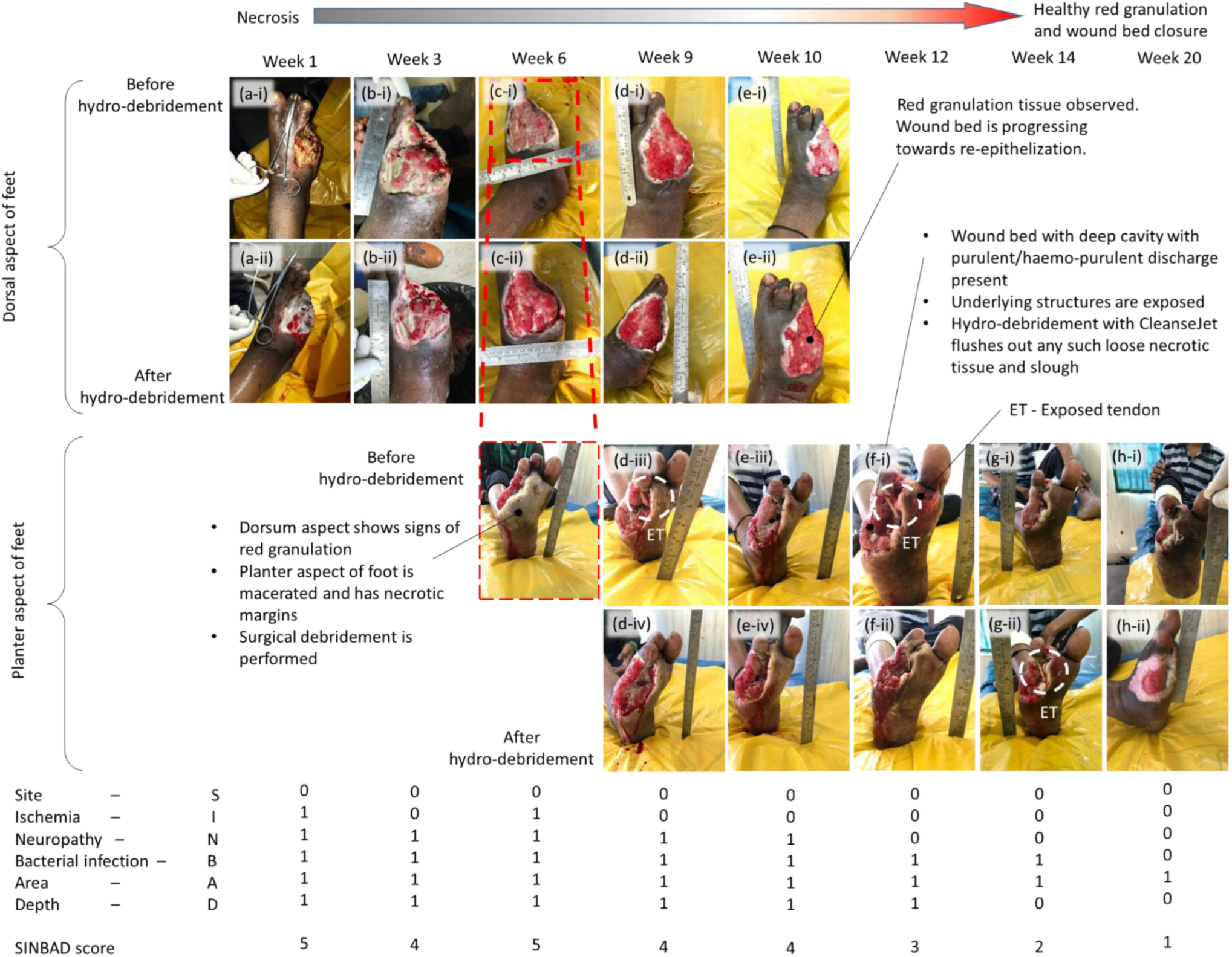
Example of a participant in his early forties with uncontrolled diabetes, presented with wet gangrene on the fourth and fifth toes of the right foot, extending to the midfoot. After surgical debridement and amputation of fourth and fifth toes, further progress of necrosis/gangrene required repeated surgical debridement. Treatment included strict diabetic control, broad-spectrum antibiotics, and weekly hydro-debridement using the CleanseJet. Irrigation with CleanseJet using modulated pressure removed any loose necrotic tissue and discharges, with negligible trauma to newly-formed granulation tissue at the wound bed. Comprehensive wound management and lavage with CleanseJet resulted in complete wound closure within 15 weeks

Progressive granulation and micro-bleeding indicated tissue regeneration following the debridement (**Figure 5** b-ii – c-ii). By the ninth week, the wound on the dorsum of the foot showed significant improvement (**Figure 5** d - i), with progressive coverage by healthy granulation tissue. However, during follow-up, the participant developed plantar cellulitis (see **Figure 5** c – ii inset), with a deep pus-filled pocket (**Figure 5** c - d), requiring additional surgical drainage and amputation of the second toe. (**Figure 5** f-i) to prevent further spread. Over the following weeks, the exposed tendon of the third toe (marked as ET in **Figure 5** d-iii, f-i, and g-ii) was gradually covered by healthy granulation (see **Figure 5** f - h), demonstrating CleanseJet’s precision and ability to preserve the vital structures. Over twenty weeks, SINBAD scores gradually improved from 5/6–4/6 for the first 10 weeks to 1/6 by the 20^th^ week, with complete granulation and significant re-epithelialization.

## Discussion: benefits and limitations

A qualitative outcome of wound management with CleanseJet, as discussed in *ESI* **S3.0**, coupled with cases discussed above and in **S4.0** in the *ESI*, highlight the viability of pressurized hydro-debridement by the CleanseJet in clearing devitalized tissue debris from complex wound-beds. A comparative analysis of observed clinical outcomes across debridement methods is presented in the following section.

### Qualitative comparison of hydro-debridement using CleanseJet with other modes of wound management

Qualitative comparison with other debridement modalities highlights potential practical advantages (see **Table 4**).

**Table 4:**
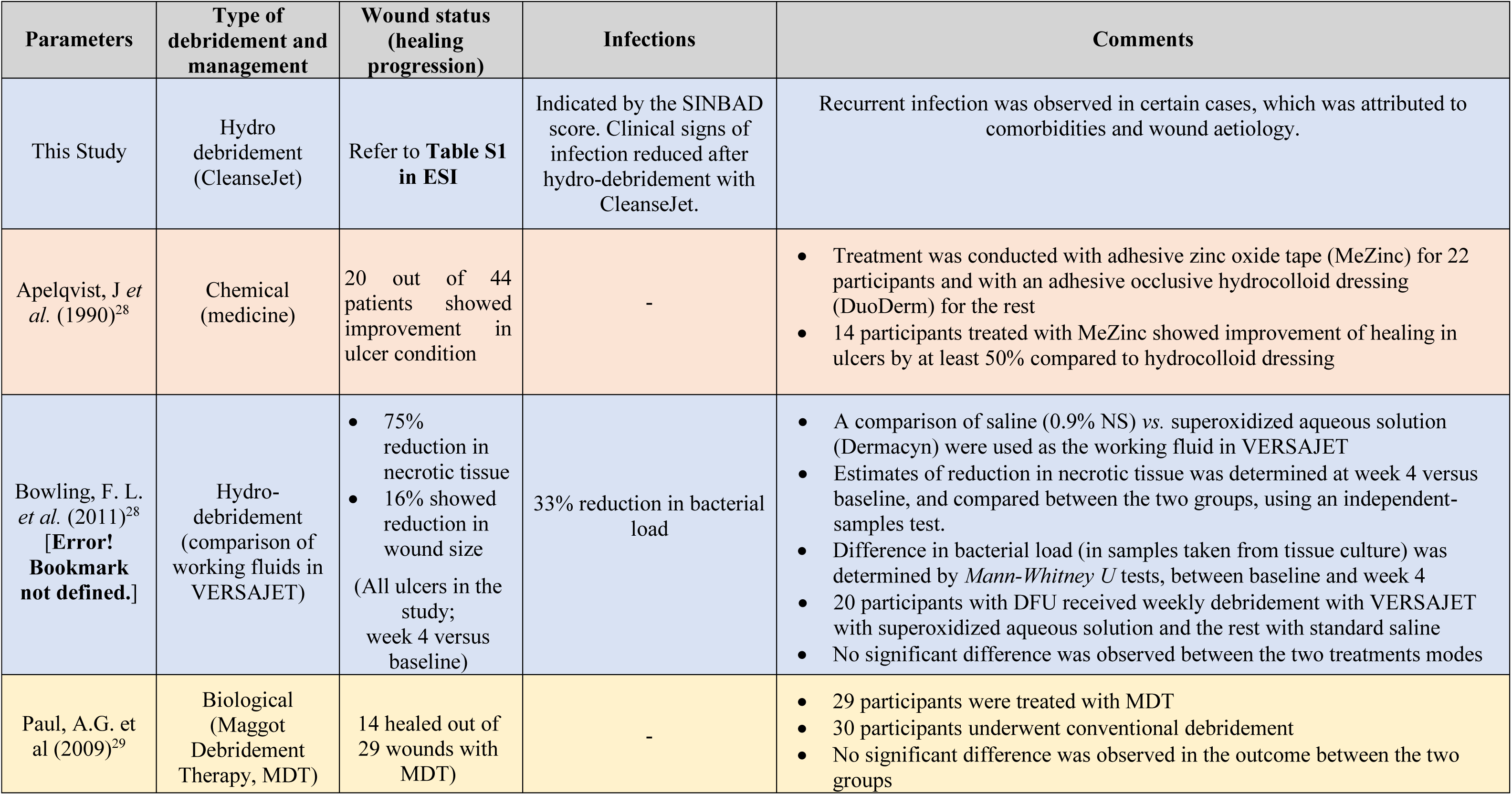
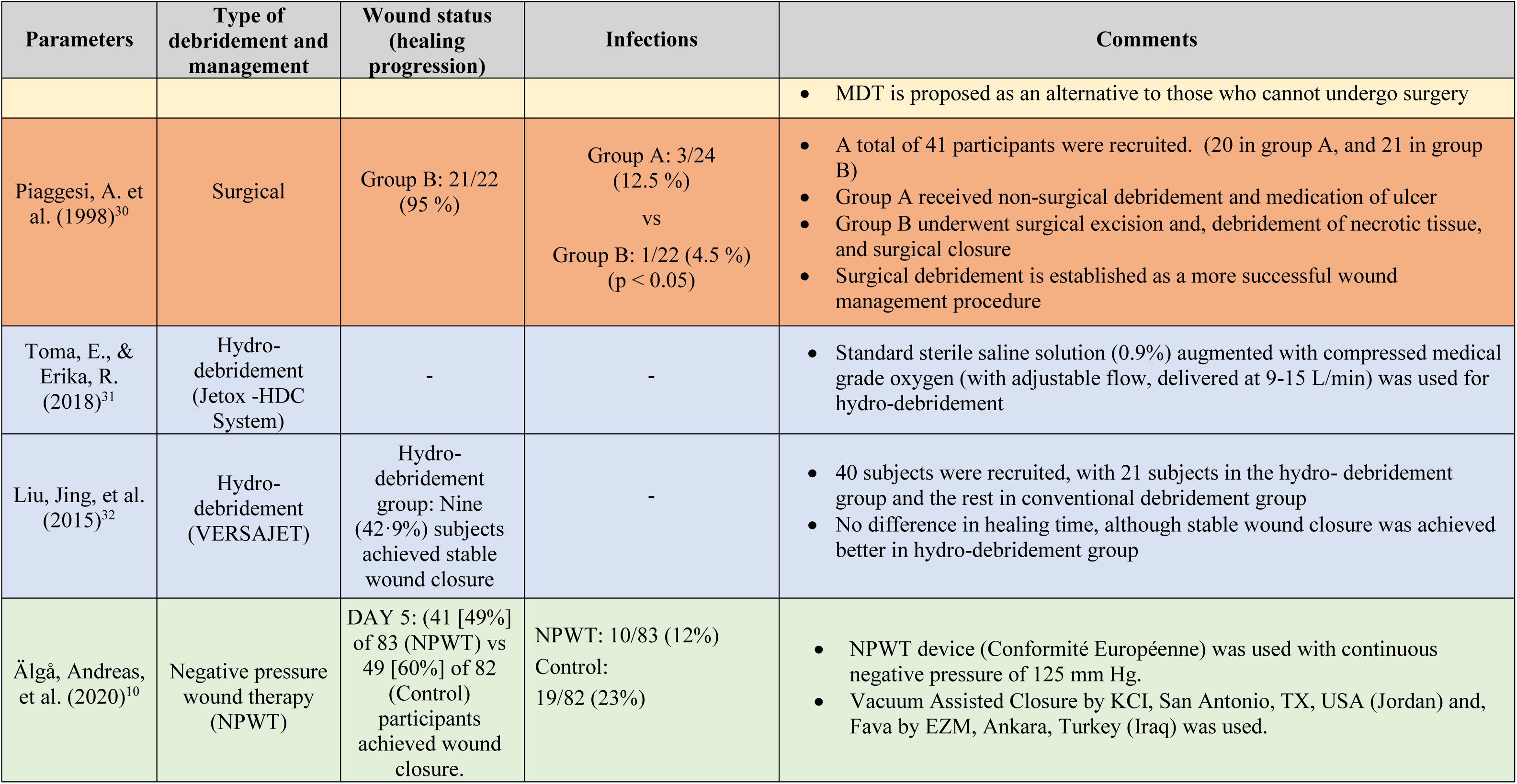
Comparison of healing and presence of infection, of hydro-debridement using CleanseJet in the present study with other debridement and management.

Healing was defined clinically by the presence of healthy granulation tissue (SINBAD score <3; **Table S1**, *ESI*) without signs of infection, while infection rate was determined by recurrence during follow-up (**Figure S4,** *ESI*). Enzymatic and autolytic methods offer selective tissue removal but are slower and may be suboptimal in wounds with heavy bioburden. NPWT is effective for complex wounds but may be constrained by cost, infrastructure requirements, and limited adaptability to anatomically irregular surfaces.^30^ In contrast, commercial hydrosurgery systems provide efficient debridement but are often associated with high capital and disposable costs.

### Innovation: Comparative assessment with other hydro-debridement tools

Key features of CleanseJet and comparable commercial hydrosurgery systems^12^ are summarized in **Table 5**. CleanseJet achieves comparable outcomes at a fraction of the cost (∼$500), with a reusable and autoclavable handpiece, reducing operating expenses substantially. Its modular design allows interchangeable nozzles of varied flow rates and spray fan angles depending on wound types, facilitating hydro-debridement of deep cavities, small apertures, and hard-to-reach areas (**Movies 3–5** – *ESI*; **Figures 4–5**, **S4** – *ESI*). Operating at ∼5 bar – far lower (∼1/100^th^) than that in the commercial devices – the device offers controlled,^20^ adjustable-pressure irrigation with simplified setup. CleanseJet improves wound visualization during procedures, maintains consistent operating costs across the wound sizes, and is suitable for outpatient and bedside use.

**Table 5:**
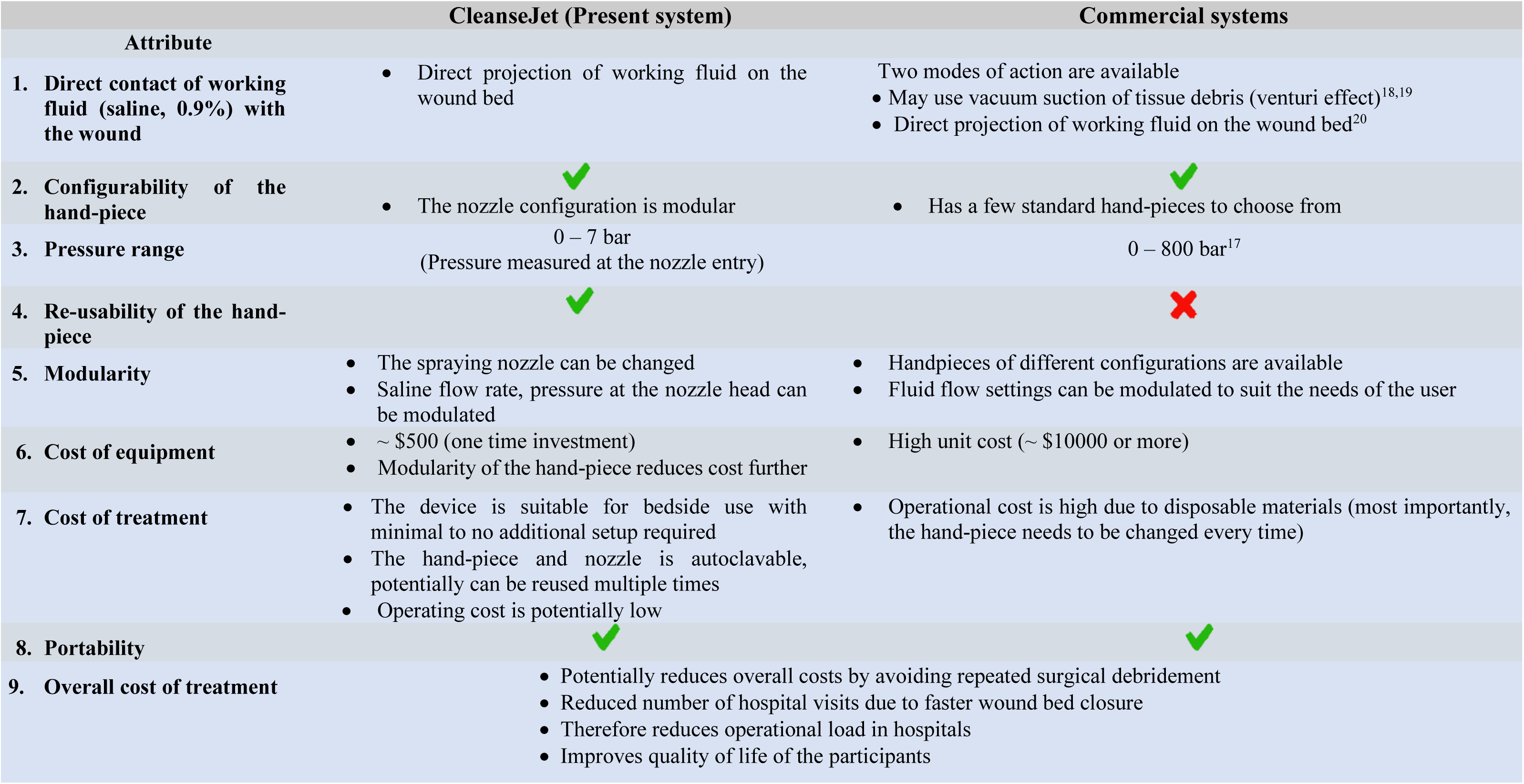
Comparing the CleanseJet with commercial hydro-debridement systems^18, 19, 20,33^.

It is important to note that the device cost of ∼US-$500 reflects only the fabrication cost of the prototype and does not account for expenses associated with research, validation and development of the prototype. With subsequent scaling-up of the device, we expect the base product cost to go down, but it is also expected that regulatory approval, distribution, marketing, clinician training, or post-market support will contribute to an increase in the price of the product. Consequently, direct cost comparisons with commercially available wound debridement systems fall beyond the scope of the present study. The objective of this work is instead to demonstrate the technical feasibility and clinical utility of a simple hydro-debridement device capable of achieving effective wound debridement, rather than to provide a comprehensive economic evaluation.

### Limitations and recommendations for future study

Similar to other hydro-debridement systems, CleanseJet is not suitable for dry necrotic eschar, which requires prior mechanical and/or surgical debridement. Nevertheless, CleanseJet’s overall performance, ease of use, and cost-effectiveness make it a valuable option for healthcare facilities as a strong alternative to existing hydro-debridement platforms. Alongside this glaring advantage, a few limitations of the current study also warrant consideration. The study was open-label, nonrandomized, and included a small comparator group, limiting statistical inference. Outcomes were largely qualitative and based on clinical assessment rather than reduction in quantitative wound area measurements. Second, wound healing outcomes were assessed using the Wagner and SINBAD scoring systems, these tools are primarily validated for diabetic foot ulcers. In the present study, the study population included a heterogeneous group of soft tissue wounds, including wounds of the hand and other anatomical sites. Additionally, heterogeneity in wound etiology and comorbidities introduces confounding variables. Larger randomized controlled trials with standardized healing endpoints and cost-effectiveness analyses are required to confirm comparative efficacy and broader clinical utility.

## Statements and declarations

### Author contributions

**Arkadeep Datta:** Writing – review & editing, Writing – original draft, Visualization, Investigation, Data curation, Formal analysis.

**Rudrajit Majumder:** Visualization, Investigation, Data curation.

**Indranil Biswas:** Data curation, Writing – review & editing

**Ranjan Ganguly:** Writing – review & editing, Supervision, Funding, Project administration, Methodology, Formal analysis, Conceptualization.

**Apurba Kumar Santra:** Funding, Conceptualization, Supervision

**Manas Kumar Gumta:** Funding, Conceptualization, Resources, Writing – review & editing, Formal analysis.

**Subhasish Sarkar:** Conceptualization, Supervision, Resources, Project administration, Funding acquisition, Formal analysis.

## Supporting information

ESI document

## Data Availability

Data and materials availability: All the data, codes, and materials are kept in the Department of General Surgery, College of Medicine Sagore Dutta Hospital. These are available to any researcher after approval from the authority.

## Acknowledgements

The authors are thankful to Dr. Rohan Sinha, senior resident physician, CMSDH, for their valuable insights and involvement that contributed to this work. The authors are grateful to Mr. Sanjib Biswas, LDC; paramedics Mr. Md Jahid Ansari and Mr. Amit Biswas of CMSDH for the assistance.

## Ethics approval

Ethical approval (Memo No: - CMSDH/IEC/256/11-2021 Dated: - 15/11/21) to perform the study was obtained from the Institutional Ethical Committee of the College of Medicine Sagore Dutta Hospital, with registration number: - ECR/1210/INST/WB/2019. Research was conducted according to the ‘*National Ethical Guidelines for Biomedical and Health Research Involving Human Participants’* by the Indian Council of Medical Research, India.

## Consent to participate

All the participants included in the study were properly counseled and signed the informed consent form, as approved by the IEC. The manuscript does not contain any identifiable images or personal data of participants.

## Consent to Publish

Informed written consent to publish was obtained from all participants before participation in the study, with approval from the Institutional Ethics Committee. All images have been anonymized, and no personal identifiers are disclosed.

## Data and materials availability

All the data, codes, and materials are kept in the Department of General Surgery, College of Medicine and Sagore Dutta Hospital. These are available to any researcher after approvals through appropriate channels.

