## Supplementary material for "A Tunable Flat-Jet Hydro-Debridement Device: Clinical Feasibility for Soft Tissue Wound Management": ESI document

---

Current address:
* INPHYNI - Institut de Physique de Nice, Université Côte d'Azur – CNRS, Nice, France
[†] Department of A*erospace and Mechanical Engineering,* University of Southern California, USA

### S1.0 Wound Classification

#### The Wagner grading

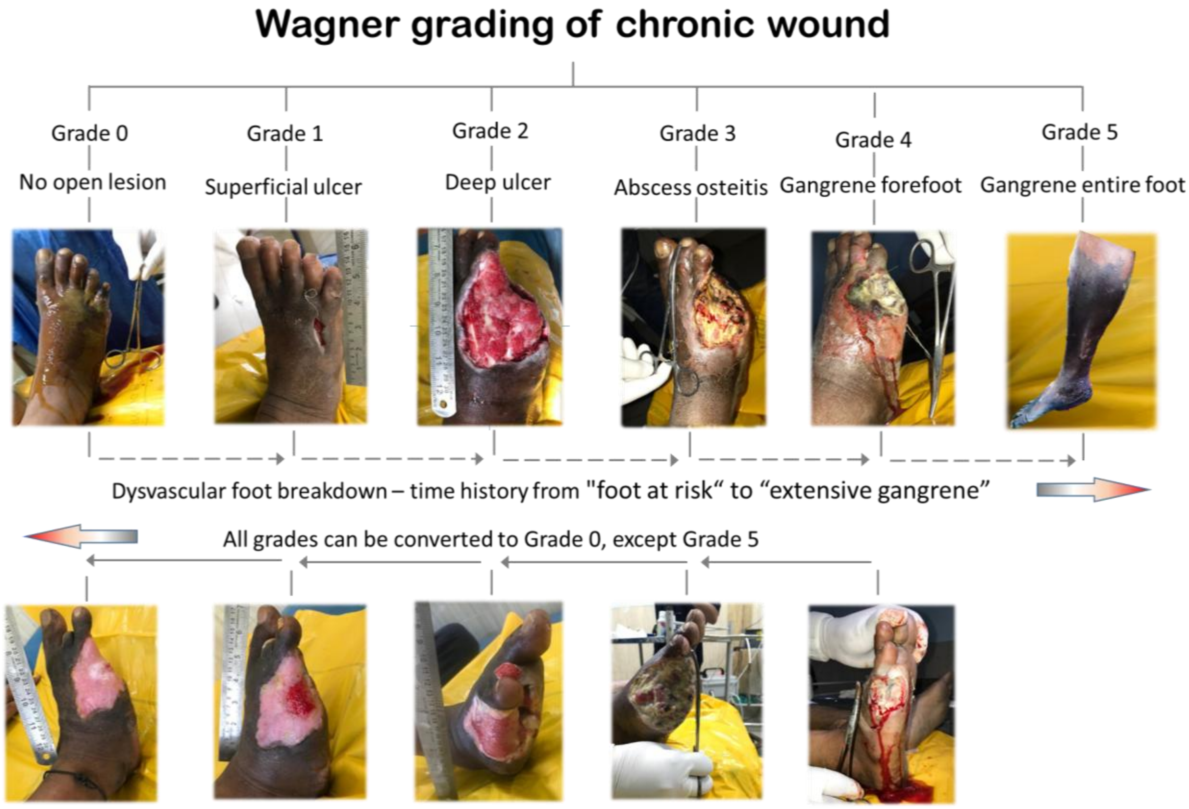


**Figure S1:** The Wagner grading[1] system is used to stage the ulcers in patients upon their admission to the ward. This system facilitates a rapid assessment of wound severity, enabling prompt prescription of appropriate wound management protocols.

### S2.0 Operational overview of the CleanseJet system: A scenario-based comparison 'before' and 'after' application

**Figure S2 A-i** shows the wound in a participant with a diabetic foot ulcer (DFU) with amputation of the fourth and fifth toes (discussed in detail Case 3 in the main text). The wound is presented with a necrotic pale yellow biofilm layer covering both hard (exposed bones) and soft tissues (muscle tissue, exposed tendons and fascia). Debriding such thin biofilms following the SoC risk damaging the underlying tissues. In contrast, CleanseJet uses a stream of normal saline projected onto the wound-bed under controlled pressure, visibly removing loose necrotic tissue and biofilm and revealing healthy fascia underneath. Mobility of the adjoining digits indicates no damage to the exposed tendons or muscles (see **Figure S2 A-ii**). Some micro-bleeding observed during the procedure indicates stimulated blood circulation in the region.

During follow-up visits (**Figure S2 B-i**), the same participant seemed to exhibit significant healing with granulation tissue covering the previously exposed tendons and bones (see **Figure S2 A-ii**). However, this granulation is seen covered with whitish biofilm and heavy fibrin deposits, accompanied by some serous discharge. **Figure S2 B-ii** shows the exposed healthy granulation tissue after the pressurized saline spray from the CleanseJet have dislodged this unwarranted loose necrosis. The spray flowrate and pressure are modulated such that only the biofilm is removed, while the underlying would bed remains unharmed.

In a more severe scenario (**Figure S2 C-i**), the participant is seen to have developed a purulent cavity at the footpad. This, along with a foul-smelling discharge – indicative of a serious infection likely exacerbated by the accompanying health issues and poor diabetic control – appeared to have worsened the wound condition. As previously discussed, conventional chronic wound management would be ineffective in complete removal of slough tissue from the cavities in such complex wounds. Additionally, traditional mechanical methods can inflict cause trauma, particularly in cases where such exposed tendons are visible, further delaying wound closure. CleanseJet provides a targeted solution with adjustable flow and spray pressure, effectively removing the devitalized tissue while preserving any tendons, soft tissues, and neo-vessels that may be present (see **Figure S2 C-ii**).

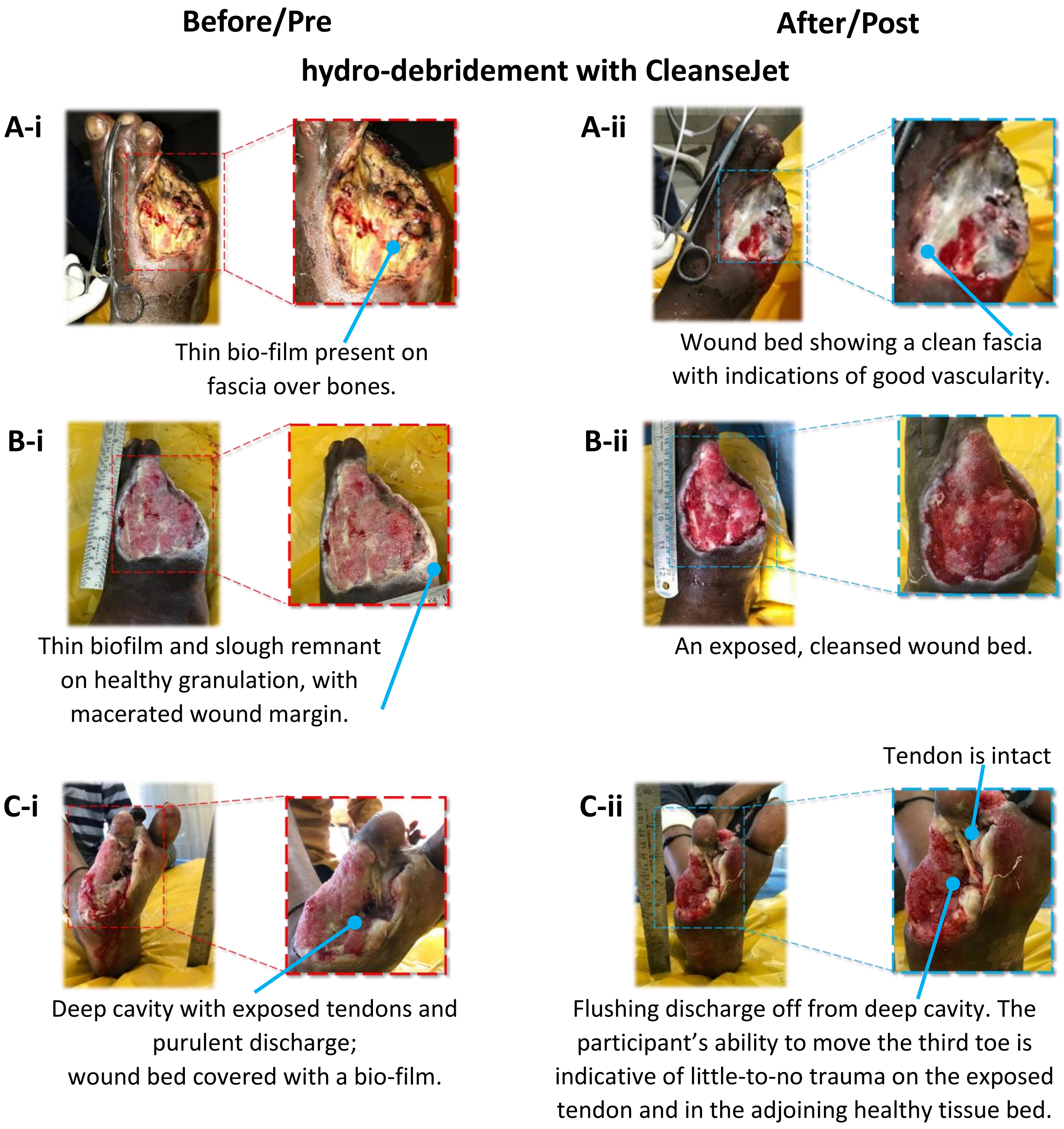


**Figure S2** Hydro-debridement in a chronic wound using the CleanseJet.
**(A – i)** A participant with a diabetic foot ulcer (DFU) and amputated fourth and fifth digits on the right foot underwent surgical debridement to remove black necrotic tissue on the dorsum, that extended to the midfoot. A yellowish layer of biofilm is seen to conceal the fascia covering both hard (bone) and soft (muscle, tendon) tissues. **(A – ii)** Irrigation with CleanseJet removes all visible biofilm, exposing the white fascia underneath. The presence of micro-bleeding indicates the vascularity in the wound bed to be intact.

**(B – i)** The participant presents with an inflamed right foot with DFU, extensive slough, and biofilm deposits on healthy granulation tissue. **(B – ii)** CleanseJet irrigation significantly removes devitalized tissue, exposing healthy granulation underneath.

**(C – i)** The wound bed (DFU) is observed to have progressed to the planter aspect of the foot, with the presence of a deep cavity filled with purulent discharge. Biofilm and slough tissue are present on the adjoining granulation tissue and a tendon attached to the third toe is seen to be exposed. **(C – ii)** Post hydro-debridement, the wound bed is seen to be clear of the devitalized tissues; the deep cavity is flushed out of any discharge. Ability of the participant to move the third toe indicates the tendon to be intact and functional.

### S3.0 Qualitative outcome of wound management with CleanseJet

This pilot evaluation suggests that pressurized hydro-debridement using CleanseJet is clinically feasible for removal of devitalized tissue from complex wound beds. Some studies[5] consider the complete closure of the wound bed as the healing target, while others emphasize more on the rate of wound bed closure[6] as a meaningful metric – both influenced by wound characteristics, comorbidities, and infection control strategies. In the absence of a universally accepted definition of "time-to-heal," outcomes in this study were defined by the emergence of healthy red granulation tissue, a pragmatic and clinically meaningful indicator of wound bed readiness. **Figure S3** shows participant healing timelines using SINBAD scores. The colour map of each tile in **Figure S3** designates the progression of healing in the wound beds – from greyish (denoting presence of necrotic tissues) to red (generation of red granulation on wound) – albeit it being qualitative in nature. Serial SINBAD scoring demonstrated progressive transition from necrotic or slough-covered tissue to granulating wound beds. Infection control was assessed using established clinical indicators, including reduction in exudate, erythema, edema, malodour, pain, and visible granulation – which are well-accepted surrogate markers of infection control in wound care[7]. Although wounds with similar baseline SINBAD scores were expected to follow comparable trajectories, healing times varied, underscoring the influence of wound size, depth, neuropathy, glycemic status, and infection recurrence. Larger and neuropathic wounds showed delayed granulation, whereas superficial wounds without significant comorbidities progressed more rapidly. For instance, wound beds 1 and 2 had delayed emergence of healthy granulation, largely due to extensive wound size, depth, and presence of neuropathy due to diabetes, while wound-beds 10 and 13 demonstrated faster recovery. Despite having a relatively high SINBAD score and a large wound, participant 14 demonstrated a rapid healing process. This may be attributed to the superficial nature of the wound and the absence of comorbidities.

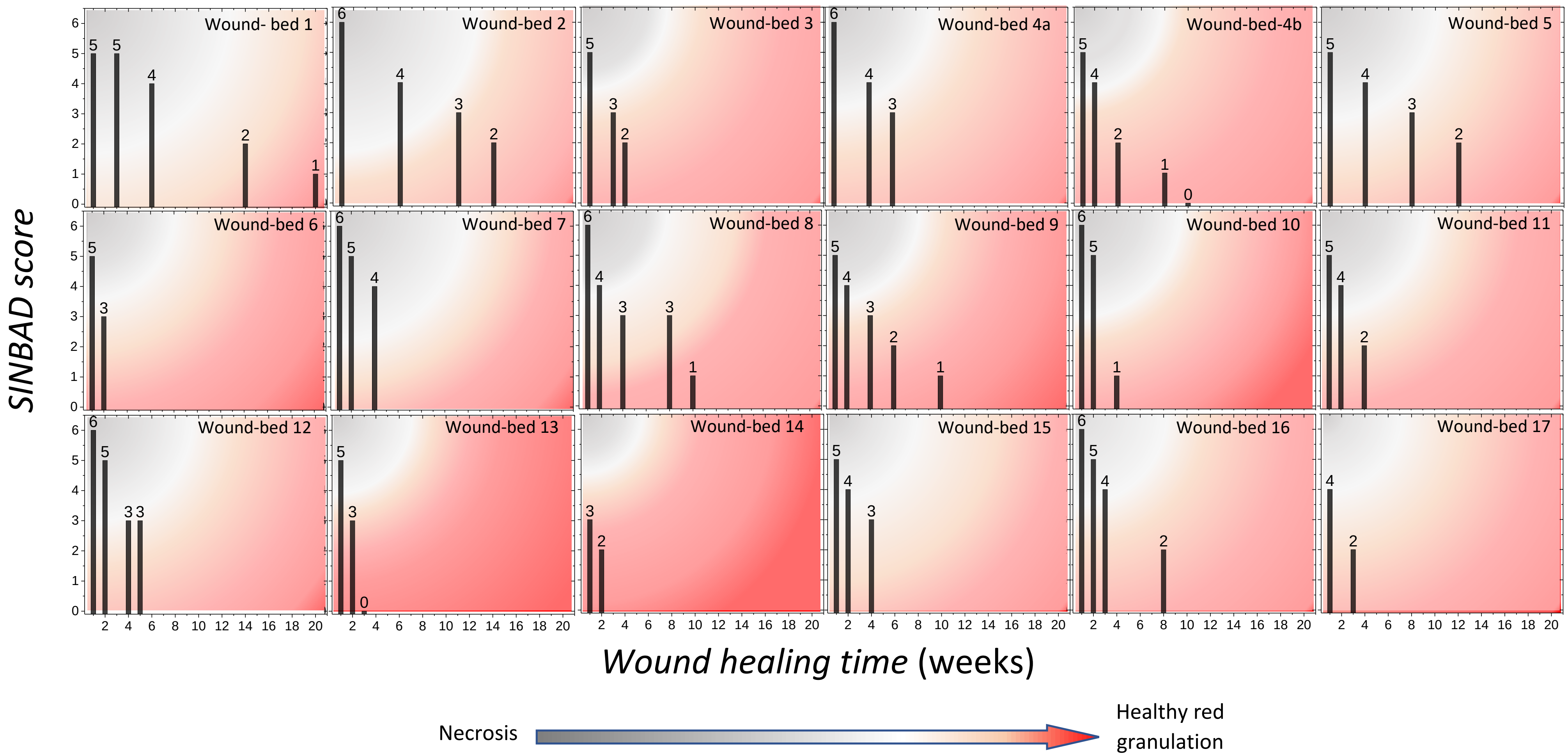


**Figure S3:** Wound healing timeline, quantified using the SINBAD score, for 17 participants (wound-bed 1 through 17, participant - 4 having two wound-beds a and b). Poor glycemic control, other comorbidities, large wound size, age are some of the factors leading to delayed wound healing.

### S4.0 Regular tracking of wound in participants with SINBAD classification and qualitative assessment of wound healing time

A total of 19 wounds sites in 20 participants are tracked in the study (including 17, and their weekly wound progression is depicted in the **Figure S4**, with the time taken for each wound to exhibit healthy granulation being noted in **Table S1**. While the wound-bed of the participants are stratified using WAGNER grades at their initial report, the progress of wound healing is qualitatively described using the SINBAD score. The participant for wounds 4a and 4b is the same, but due to differences in the size, shape and site of the wound bed are designated differently. Observations were classified using the SIDBAD scoring system to qualitatively assess wound healing. The weekly progression of the wound site (see **Figure S4**) reveals a marked development in reduction of infections and exudate, improved angiogenesis and healthy granulation tissue formation. It is also observed from **Table S1** that participants who received hydro-debridement with CleanseJet for at least 3 weeks exhibited improvement in the SINBAD score of their wounds. Debridement of two additional wound-beds (see the cases of wound-bed 18 and 19 in **Table S1**) were carried out when the CleanseJet was able to irrigate hard-to-reach areas and wound clefts (see *ESI* **Movies 4** and **5**). However, these two participants did not report to the facility further and hence the progression of their wound-bed healing could not be reported.

**Table S1:** Qualitative classification, time taken to heal, and outcome.

| Wagner Grade stratification of participants at their initial report | Wound bed | Medical conditions | Time taken to heal (weeks) | SINBAD score | | | | | | | | | | | | | |
|---|---|---|---|---|---|---|---|---|---|---|---|---|---|---|---|---|---|
| | | | | Initial score | | | | | | | Final score | | | | | | |
| | | | | S | I | N | B | A | D | Total | S | I | N | B | A | D | Total |
| **Grade 0** | 11 | Cellulitis | 4 | 1 | 0 | 1 | 1 | 1 | 1 | **5** | 1 | 0 | 0 | 0 | 1 | 0 | **2** |
| **Grade 1** | 2 | T2DM | 14 | 1 | 1 | 1 | 1 | 1 | 1 | **6** | 1 | 0 | 0 | 0 | 1 | 0 | **2** |
| | 3 | T2DM | 4 | 1 | 1 | 0 | 1 | 1 | 1 | **5** | 1 | 0 | 0 | 0 | 1 | 0 | **2** |
| | 5 | T2DM | 12 | 1 | 1 | 0 | 1 | 1 | 1 | **5** | 1 | 0 | 0 | 0 | 1 | 0 | **2** |
| | 14 | Burn Injury | 2 | 1 | 0 | 0 | 1 | 1 | 0 | **3** | 1 | 0 | 0 | 0 | 1 | 0 | **2** |
| | 16 (Case 1*) | T2DM (Poor glycemic control) | 8 | 1 | 1 | 1 | 1 | 1 | 1 | **6** | 1 | 0 | 0 | 0 | 1 | 0 | **2** |
| | 18 (*ESI* – Movie 4) | Cellulitis | - | 1 | 0 | 1 | 1 | 1 | 1 | **5** | The participants did not report to the facility further; therefore, the wound healing progression could not be tracked. | | | | | | |
| | 19 (*ESI* – Movie 5) | T2DM | - | 0 | 0 | 1 | 0 | 1 | 1 | **3** | | | | | | | |
| **Grade 2** | 4b | T2DM | 10 | 0 | 1 | 1 | 1 | 1 | 1 | **5** | 0 | 0 | 0 | 0 | 0 | 0 | **0** |
| | 6 | Accident trauma injury | 2 | 1 | 0 | 1 | 1 | 1 | 1 | **5** | 1 | 0 | 0 | 0 | 1 | 1 | **3** |
| | 8 (Case 2*) | CKD | 10 | 1 | 1 | 1 | 1 | 1 | 1 | **6** | 1 | 0 | 0 | 0 | 0 | 0 | **1** |
| | 12 | T2DM (Poor glycemic control) | 5 | 1 | 1 | 1 | 1 | 1 | 1 | **6** | 1 | 0 | 0 | 0 | 1 | 1 | **3** |
| **Grade 3** | 4a | T2DM | 6 | 1 | 1 | 1 | 1 | 1 | 1 | **6** | 1 | 0 | 0 | 0 | 1 | 1 | **3** |
| | 7 | Cellulitis | 4 | 1 | 1 | 1 | 1 | 1 | 1 | **6** | 1 | 0 | 0 | 1 | 1 | 1 | **4** |
| | 9 | T2DM | 10 | 0 | 1 | 1 | 1 | 1 | 1 | **5** | 0 | 0 | 0 | 0 | 0 | 1 | **1** |
| | 10 | T2DM | 4 | 0 | 1 | 1 | 1 | 1 | 1 | **6** | 0 | 0 | 1 | 0 | 0 | 0 | **1** |
| | 15 | T1DM | 4 | 0 | 1 | 1 | 1 | 1 | 1 | **5** | 0 | 1 | 0 | 0 | 1 | 1 | **3** |
| | 17 | Accident trauma injury | 3 | 1 | 0 | 0 | 1 | 1 | 1 | **4** | 1 | 0 | 0 | 0 | 1 | 0 | **2** |
| **Grade 4** | 1 (Case 3*) | T2DM (Poor glycemic control) | 20 | 0 | 1 | 1 | 1 | 1 | 1 | **5** | 0 | 0 | 0 | 0 | 1 | 0 | **1** |
| | 13 | T2DM | 3 | 0 | 1 | 1 | 1 | 1 | 1 | **5** | 0 | 0 | 0 | 0 | 0 | 0 | **0** |
| **Grade 5** | | No participants were recruited. | | | | | | | | | | | | | | | |

Legend: red – 1; green – 0

**Abbreviations**: T2DM**:** Type 2 Diabetes Mellitus; T1DM: Type 1 Diabetes Mellitus; CKD: Chronic Kidney Disease.

*Refer to **Section 5** in the main text for details.

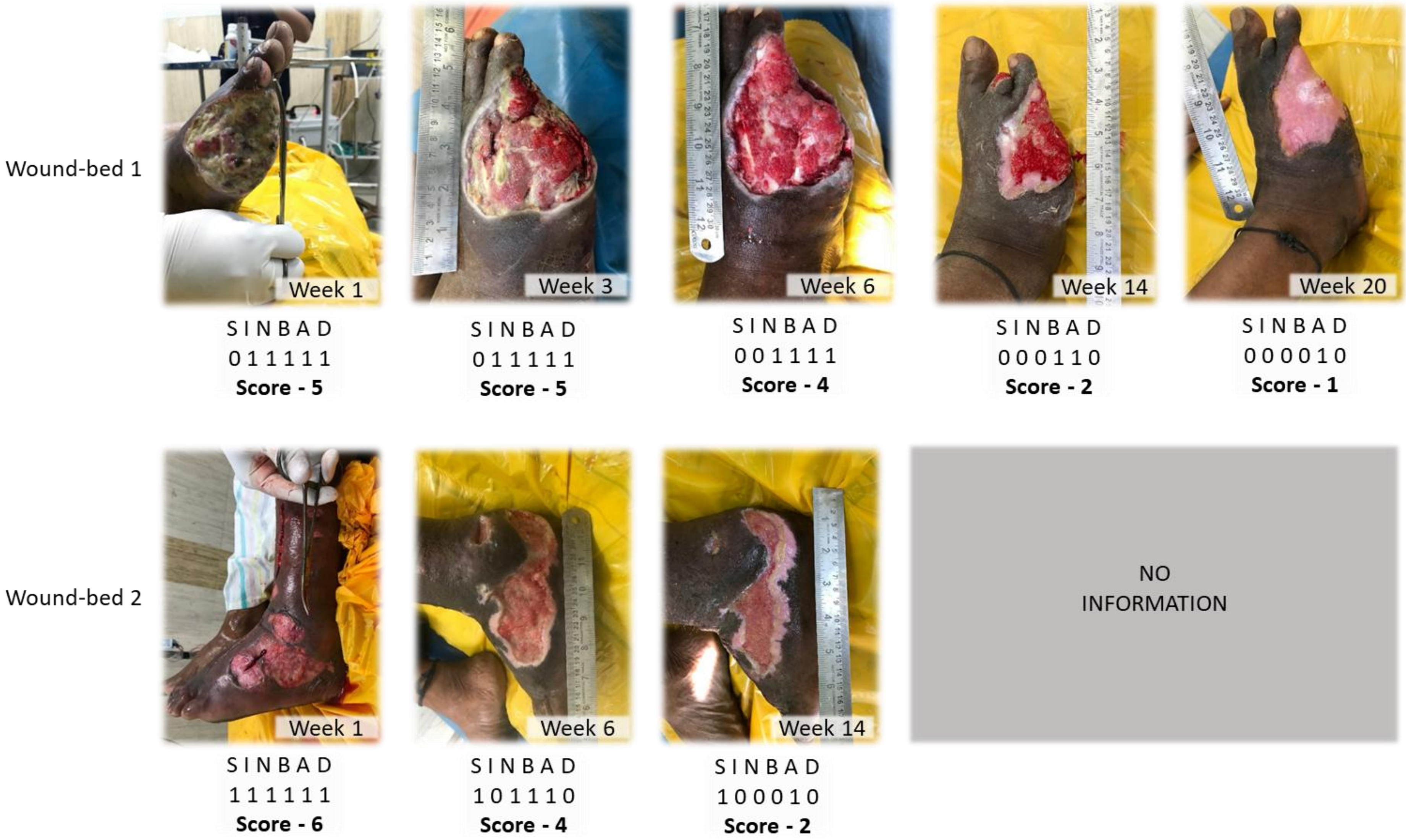
Wound-bed 1
Week 1
S I N B A D
0 1 1 1 1 1
Score - 5
Week 3
S I N B A D
0 1 1 1 1 1
Score - 5
Week 6
S I N B A D
0 0 1 1 1 1
Score - 4
Week 14
S I N B A D
0 0 0 1 1 0
Score - 2
Week 20
S I N B A D
0 0 0 0 1 0
Score - 1
Wound-bed 2
Week 1
S I N B A D
1 1 1 1 1 1
Score - 6
Week 6
S I N B A D
1 0 1 1 1 0
Score - 4
Week 14
S I N B A D
1 0 0 0 1 0
Score - 2
NO
INFORMATION

Wound-bed 3

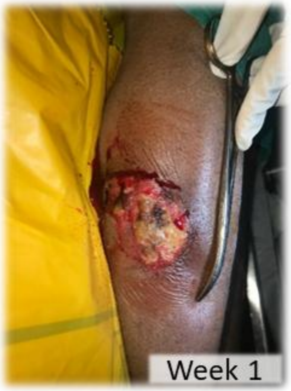


S I N B A D
1 1 0 1 1 1
**Score - 5**

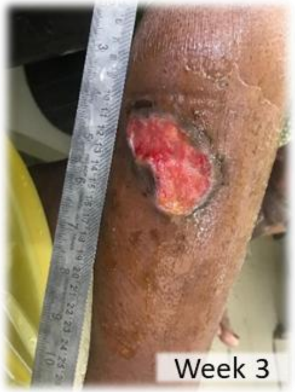


S I N B A D
1 0 0 0 1 1
**Score - 3**

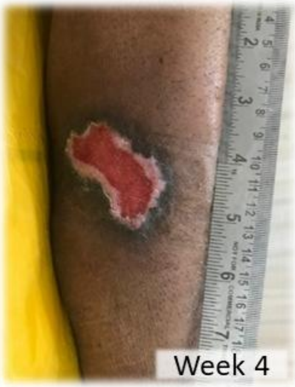


S I N B A D
1 0 0 0 1 0
**Score - 2**

NO INFORMATION

Wound-bed 4a

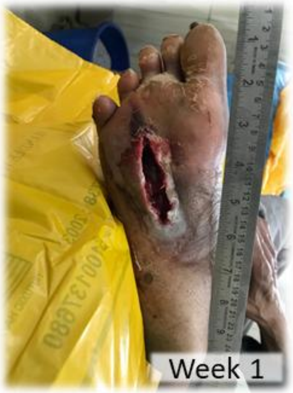


S I N B A D
1 1 1 1 1 1
**Score - 6**

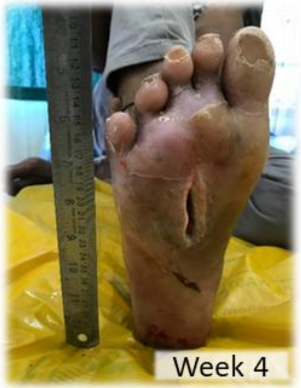


S I N B A D
1 0 0 1 1 1
**Score - 4**

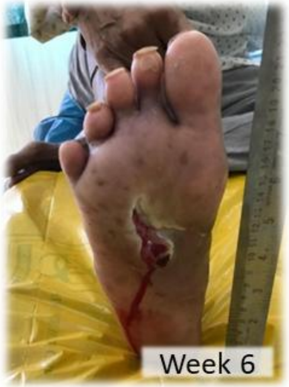


S I N B A D
1 0 0 0 1 1
**Score - 3**

NO INFORMATION

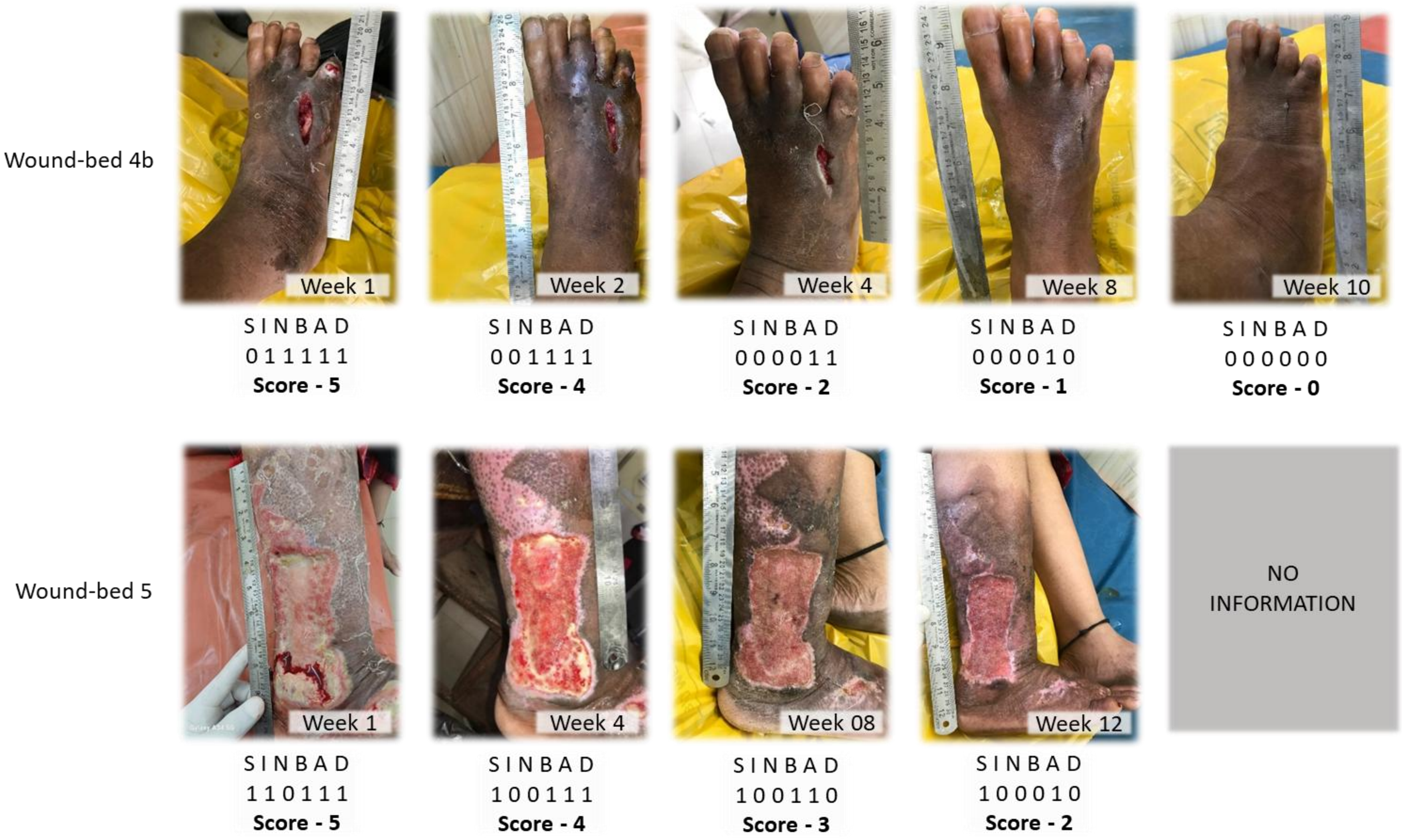

Wound-bed 4b
Week 1
S I N B A D
0 1 1 1 1 1
Score - 5
Week 2
S I N B A D
0 0 1 1 1 1
Score - 4
Week 4
S I N B A D
0 0 0 0 1 1
Score - 2
Week 8
S I N B A D
0 0 0 0 1 0
Score - 1
Week 10
S I N B A D
0 0 0 0 0 0
Score - 0
Wound-bed 5
Week 1
S I N B A D
1 1 0 1 1 1
Score - 5
Week 4
S I N B A D
1 0 0 1 1 1
Score - 4
Week 08
S I N B A D
1 0 0 1 1 0
Score - 3
Week 12
S I N B A D
1 0 0 0 1 0
Score - 2
NO INFORMATION

Wound-bed 6

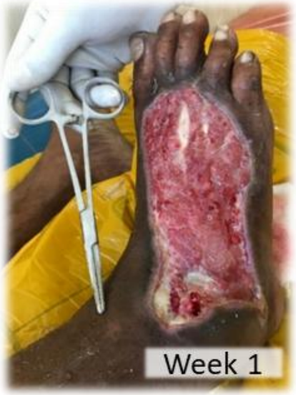


S I N B A D
1 0 1 1 1 1
**Score - 5**

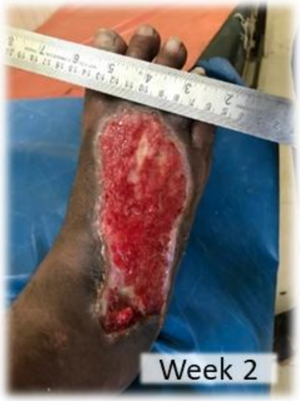


S I N B A D
1 0 0 0 1 1
**Score - 3**

NO
INFORMATION

Wound-bed 7

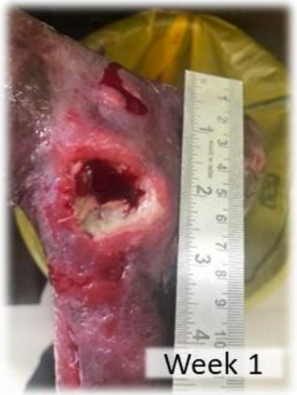


S I N B A D
1 1 1 1 1 1
**Score - 6**

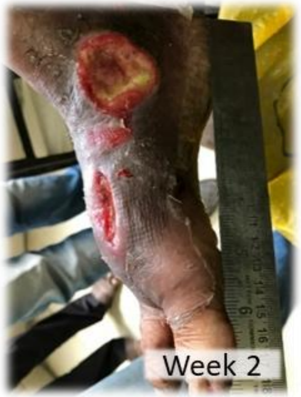


S I N B A D
1 1 0 1 1 1
**Score - 5**

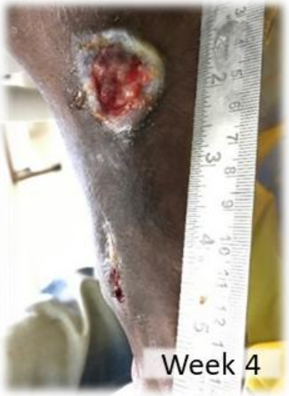


S I N B A D
1 0 0 1 1 1
**Score - 4**

NO
INFORMATION

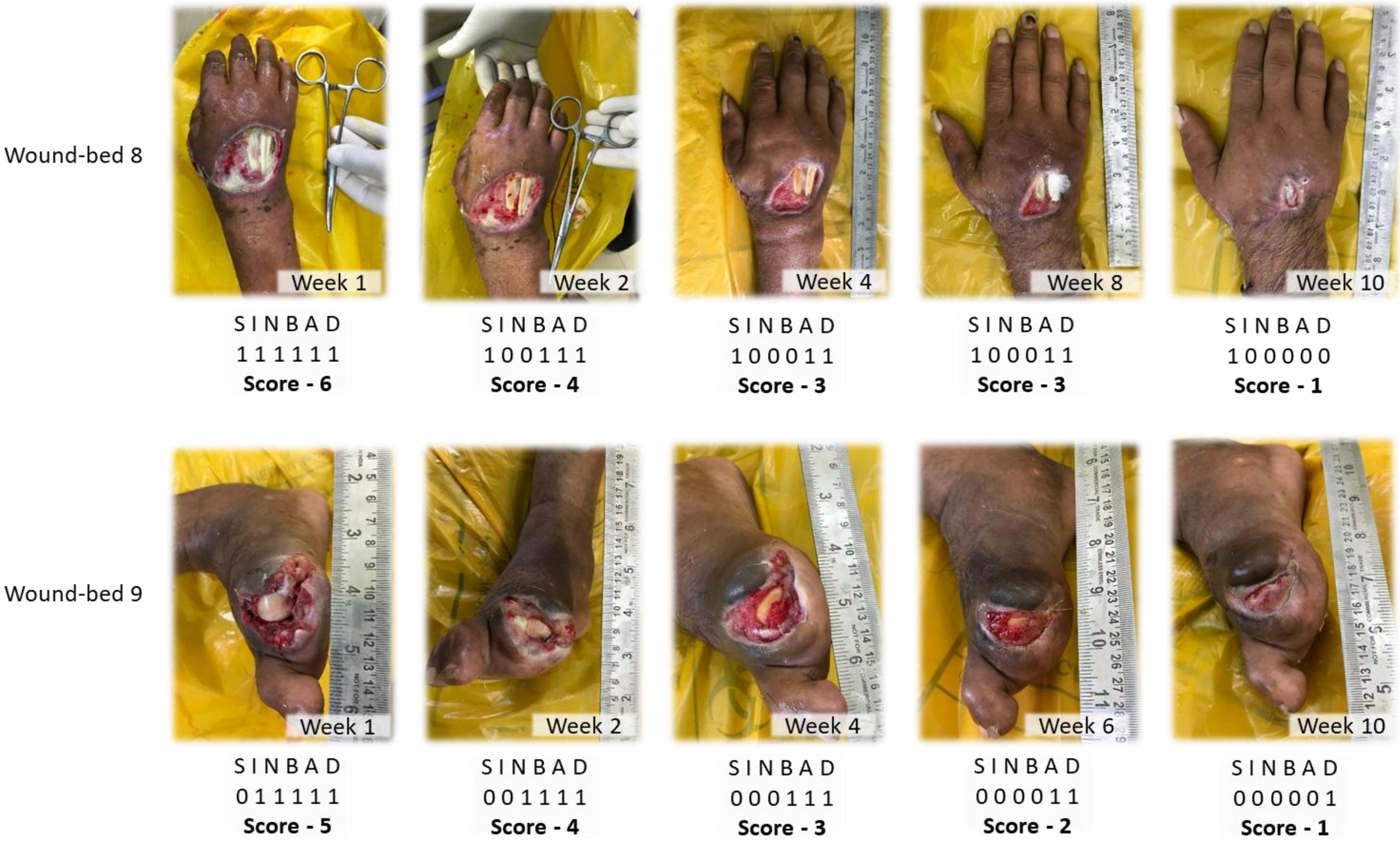
Wound-bed 8
Week 1
SINBAD
111111
Score - 6
Week 2
SINBAD
100111
Score - 4
Week 4
SINBAD
100011
Score - 3
Week 8
SINBAD
100011
Score - 3
Week 10
SINBAD
100000
Score - 1
Wound-bed 9
Week 1
SINBAD
011111
Score - 5
Week 2
SINBAD
001111
Score - 4
Week 4
SINBAD
000111
Score - 3
Week 6
SINBAD
000011
Score - 2
Week 10
SINBAD
000001
Score - 1

Wound-bed 10

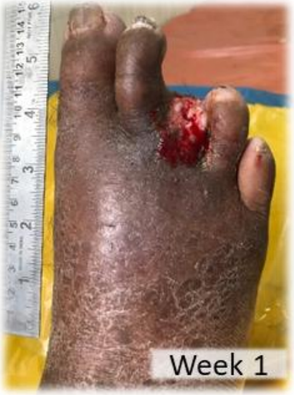


S I N B A D
1 1 1 1 1 1
**Score - 6**

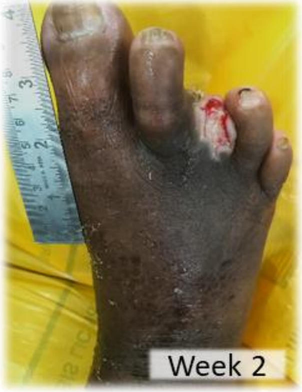


S I N B A D
1 0 1 1 1 1
**Score - 5**

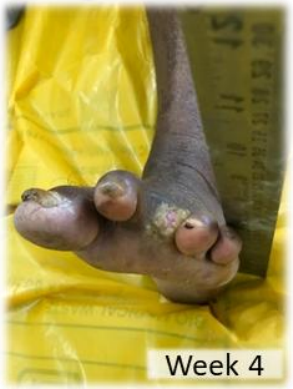


S I N B A D
1 0 0 0 0 0
**Score - 1**

NO
INFORMATION

Wound-bed 11

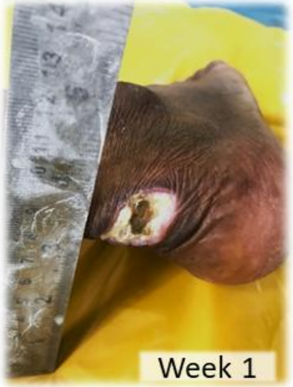


S I N B A D
0 1 1 1 1 1
**Score - 5**

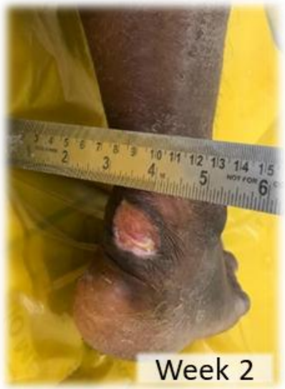


S I N B A D
0 1 0 1 1 1
**Score - 4**

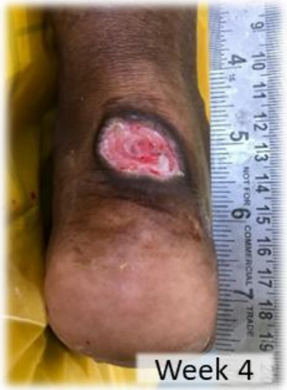


S I N B A D
0 0 0 0 1 1
**Score - 2**

NO
INFORMATION

Wound-bed 12

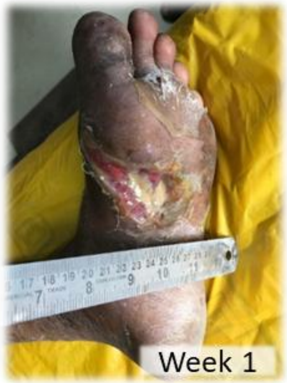


S I N B A D
1 1 1 1 1 1
**Score - 6**

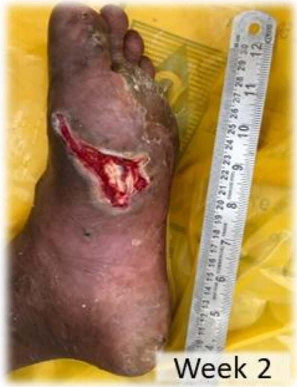


S I N B A D
1 0 1 1 1 1
**Score - 5**

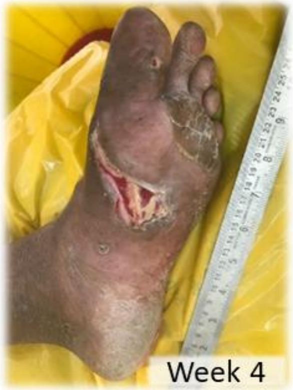


S I N B A D
1 0 0 0 1 1
**Score - 3**

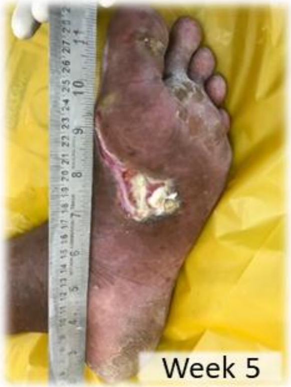


S I N B A D
1 0 0 0 1 1
**Score - 3**

NO INFORMATION

Wound-bed 13

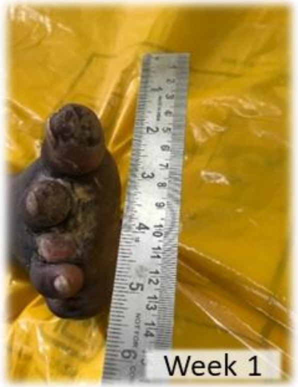


S I N B A D
0 1 1 1 1 1
**Score - 5**

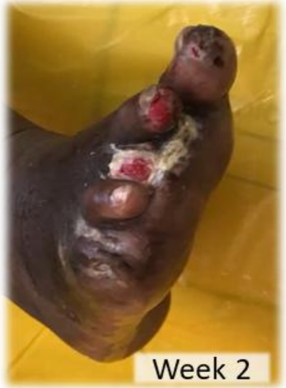


S I N B A D
0 0 0 1 1 1
**Score - 3**

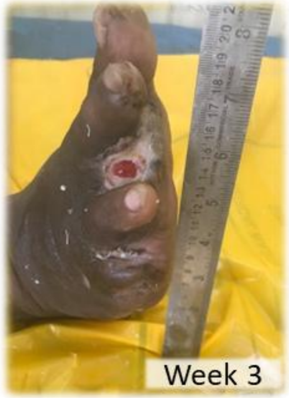


S I N B A D
0 0 0 0 0 0
**Score - 0**

NO INFORMATION

Wound-bed 14

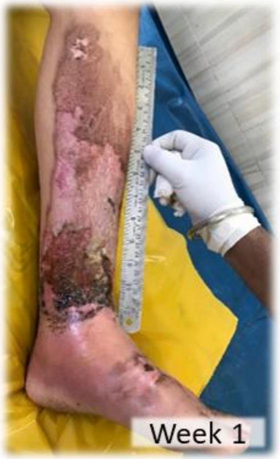


S I N B A D
1 0 0 1 1 0
**Score - 3**

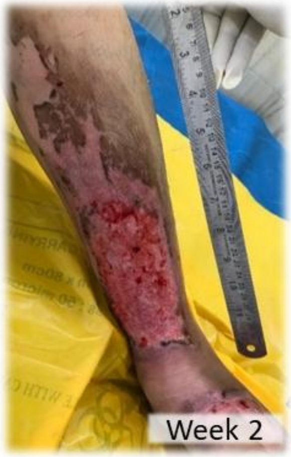


S I N B A D
1 0 0 0 1 0
**Score - 2**

NO
INFORMATION

Wound-bed 15

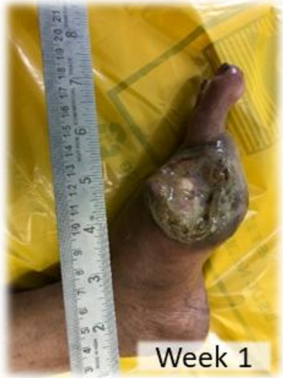


S I N B A D
0 1 1 1 1 1
**Score - 5**

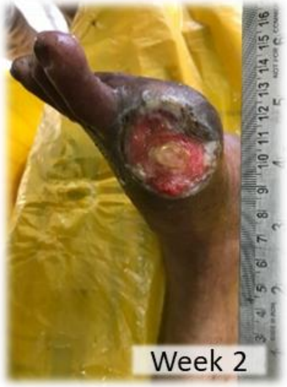


S I N B A D
0 1 0 1 1 1
**Score - 4**

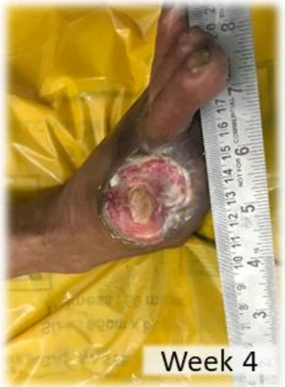


S I N B A D
0 1 0 0 1 1
**Score - 3**

NO
INFORMATION

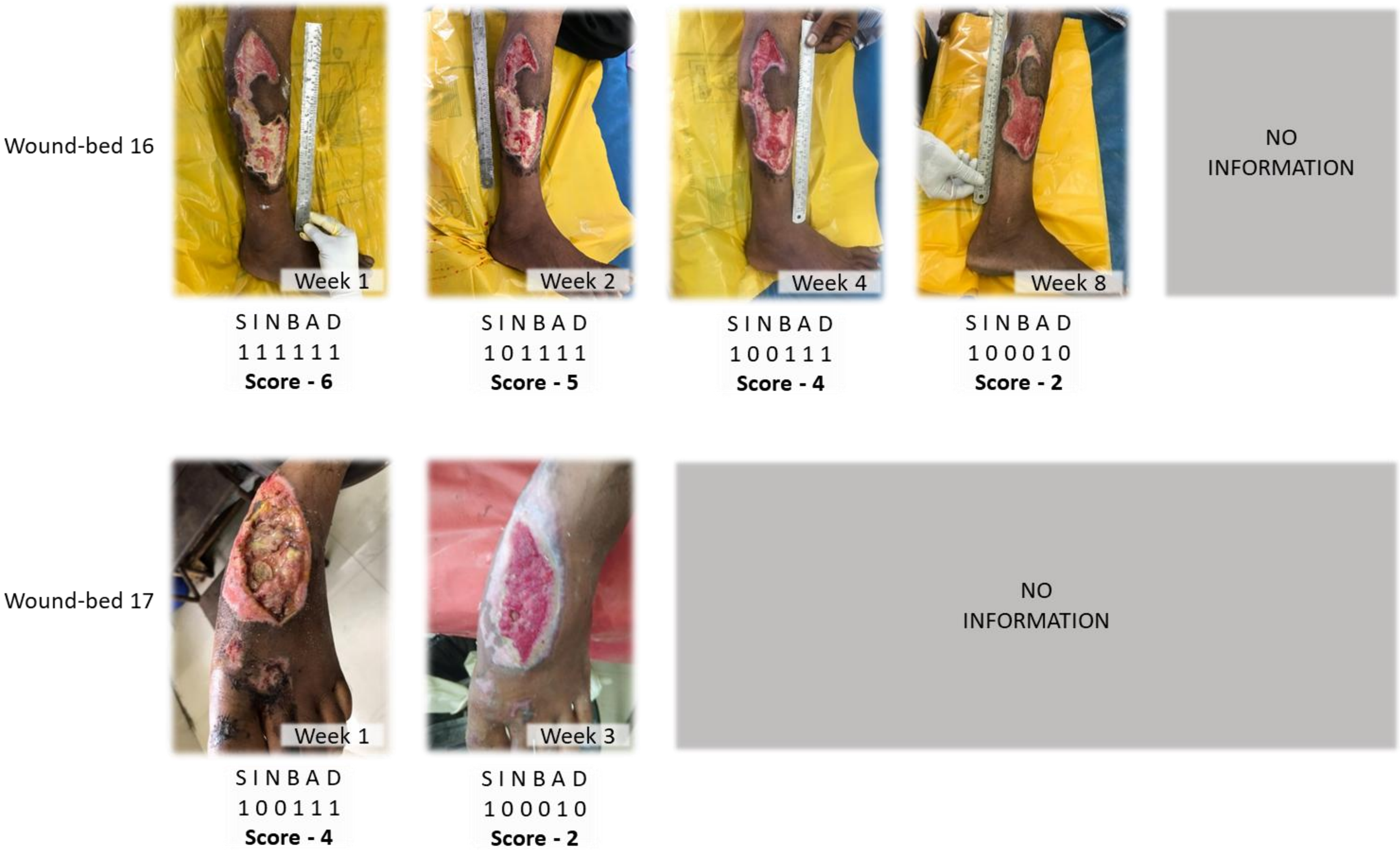


**Figure S4:** Regular tracking of wound progression, timeline, and outcomes of hydro-debridement using CleanseJet in participants is classified by SINBAD score. Tabs marked 'no information' indicate the periods when the participants were advised for home care or did not report.
